# A non-coding variant at 2p24.2 confers susceptibility to non-syndromic cleft lip and palate through LLPS-dependent regulation of *MYCN*

**DOI:** 10.64898/2026.04.07.26350283

**Authors:** Zhaoyi Wu, Zhiying Yuan, Ruihuan Yang, Zhuo Huang, Yiwei Liu, Liangdan Sun, Zhuan Bian, Miao He

**Affiliations:** State Key Laboratory of Oral & Maxillofacial Reconstruction and Regeneration, Key Laboratory of Oral Biomedicine Ministry of Education, Hubei Key Laboratory of Stomatology, School & Hospital of Stomatology, Wuhan University, Wuhan, Hubei Province, 430079, China; North China University of Science and Technology, Tangshan, Hebei Province, 063210, China

## Abstract

Non-syndromic cleft lip and palate (NSCLP) represents the most prevalent and clinically severe subtype within non-syndromic orofacial cleft (NSOFC), and 2p24.2 is the most significant reported risk locus for NSCLP. However, the causal variant at 2p24.2 and the underlying pathogenic mechanism remain unclear, limiting clinical translation. Here, we defined a 104-kb linkage disequilibrium (LD) block tagged by the lead SNP rs7552 at 2p24.2. Through a two-stage genetic screen within this block, including targeted sequencing and replication involving 2,437 Chinese NSCLP patients and 2,391 unaffected individuals, we identified a common non-coding single-nucleotide polymorphism, rs4263114, at 2p24.2 as the causal variant that confers susceptibility to NSCLP by residing within a previously unrecognized enhancer. Mechanistically, this enhancer physically bridges to the *MYCN* promoter through distal spatial contact, implicating *MYCN* as the pathogenic gene at this locus. Specifically, the rs4263114 risk variant reduces the recruitment of FOXP2 to the enhancer and disrupts liquid-liquid phase separation (LLPS)-driven droplet assembly. This biophysical defect impairs *MYCN* transcriptional activation and subsequently suppresses cranial neural crest cell (cNCC) differentiation. Notably, *MYCN* expression in cNCCs carrying homozygous risk alleles were partially restored by promoting FOXP2 LLPS. Collectively, our study functionally annotates the 2p24.2 locus and identifies a mechanism by which a non-coding variant disrupts transcription factor phase separation to increase susceptibility to NSCLP, providing a basis for future clinical translation.

**Graphic Abstract:** 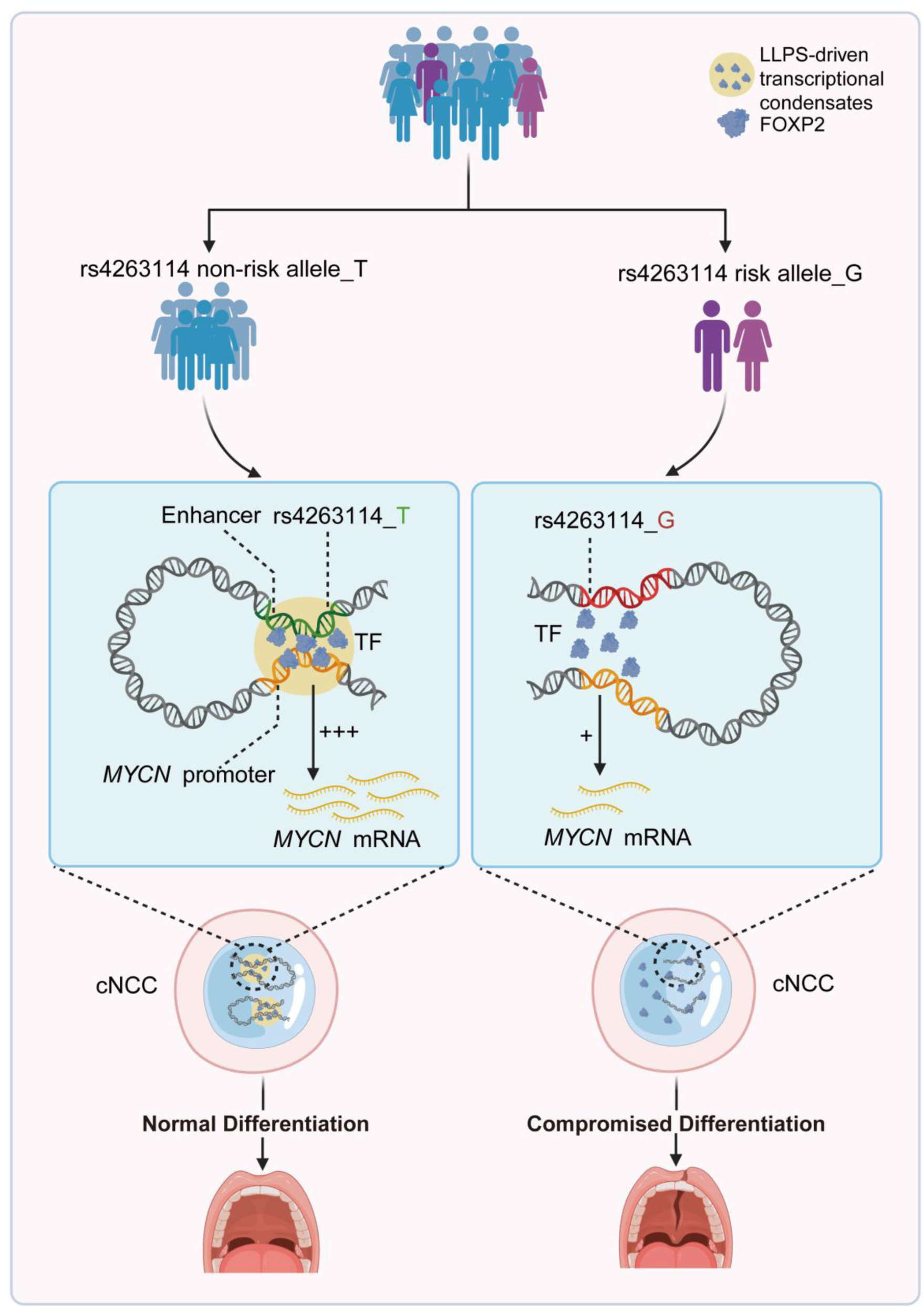

Proposed regulatory mechanism of MYCN modulated by Enh-*MYCN* across distinct rs4263114 genotypic backgrounds.

## Introduction

Among congenital craniofacial defects, orofacial clefts (OFCs) represent the most frequent manifestation, affecting approximately 1 in 700 neonates globally with a higher incidence of about 1 in 500 in East Asian and Native American populations.^1–3^ OFCs typically encompass three primary categories: cleft palate only (CPO [MIM: 119540]), cleft lip and palate (CLP), and cleft lip only (CLO). Because of their shared embryologic origin, CLO and CLP are traditionally classified as cleft lip with or without palate (CL/P [MIM: 119530]).^4,5^ Among these subtypes, CLP is the most common and severe.^6^ Distinct from the Mendelian-inherited syndromic types, non-syndromic (NS) OFC stands as the dominant form of OFC, accounting for nearly 70% of clinical presentations. These NSOFC occurrences result from intricate polygenic and environmental contributions whose hereditary landscape remains unclear.^2,7^

Following their emergence from the dorsal neural tube, cranial neural crest cells (cNCCs) migrate to the pharyngeal arches, where they differentiate into most orofacial tissues and structures.^8–10^ Precise fate determination before migration and proper regulation of cellular behavior after migration are essential for craniofacial development. Disruption of these processes is a major cause of craniofacial malformations, including OFCs.^11^ Therefore, identifying the genetic factors that regulate cNCC fate determination and cellular dynamics is essential for clarifying the pathogenesis and genetic basis of OFCs.

Since the early 2000s, large-scale genomic explorations have highlighted a diverse array of genetic contributors to NSOFC, ranging from single-nucleotide variants (SNVs), short insertions and deletions (indels), to larger copy number variations (CNVs).^12–20^ In addition, genome-wide association studies (GWAS) have successfully mapped hundreds of common variants across dozens of susceptibility loci.^21–28^ However, a major bottleneck is that the vast majority (>80%) of these risk variants are situated within non-coding intervals without functional annotation. This gap between genetic association and biological function remains a major challenge in defining the molecular mechanisms that underlie NSOFC susceptibility.^29–31^

According to the NHGRI-EBI GWAS Catalog, 2p24.2 is the top-ranked risk locus for NSCLP. However, the functional landscape in this region remains poorly defined. Specifically, it is unclear whether the reported tag SNP rs7552 at 2p24.2 is itself functional or whether other causal variants drive the association signal. Previous studies have implicated two candidate causal genes in this region: *MYCN* (*MYCN* [MIM: 164840]) and *DDX1* (*DDX1* [MIM: 601257]).^32,33^ Using a conditional knockout mouse model, our group showed that *Mycn* dosage insufficiency in cNCCs causes abnormal differentiation and hypoplasia of Meckel’s cartilage, which then disrupts tongue morphogenesis and results in palatal defects.^32^ However, our direct sequencing of the *MYCN* coding region in NSCLP families indicated that coding variants are rare.^34^ This discrepancy strongly suggests that genetic susceptibility at 2p24.2 may be driven by functional non-coding variants within cis-regulatory elements (CREs) that regulate *MYCN* expression. Meanwhile, Bartusel et al. identified a novel enhancer, e2p24.2, that regulates cNCC migration through *DDX1*-mediated tRNA splicing and thereby contributes to craniofacial morphogenesis.^33^ Thus, a key unresolved question is which causal gene is regulated by the functional variants at 2p24.2 and through what mechanism.

Here, we addressed these questions by combining genetic analysis in a Chinese NSCLP cohort with comprehensive functional annotation. We identified rs4263114 as the causal variant at 2p24.2, located within a distal enhancer. Using three-dimensional genome analysis, we demonstrated that this enhancer distally engages the *MYCN* promoter by establishing a specific long-distance chromatin interaction, supporting *MYCN* as the major effector gene. Mechanistically, we found that the rs4263114 susceptibility variant weakens the recruitment of transcription factor (TF) FOXP2 (*FOXP2* [MIM: 605317]) and impairs its liquid-liquid phase separation (LLPS) capacity. This biophysical defect reduces *MYCN* transcription, leading to impaired cNCC differentiation and craniofacial malformations, a phenotype recapitulated in our human organoid models. Collectively, our study functionally annotates the major GWAS locus 2p24.2 and identifies a new etiologic mechanism in which a non-coding variant perturbs TF phase separation to drive susceptibility to OFCs.

## Methods

### Targeted Sequencing, Genotyping and SNP Selection

Individuals with NSCLP and population-based controls were recruited from the Han Chinese population. Following formal approval from the Ethics Committee of the School of Stomatology at Wuhan University (Ethics Approval No. 2016-14), written informed consent was gathered from all individuals involved or their legal guardians. Affected individuals were diagnosed with NSCLP on the basis of physical examination.

Based on prior GWAS results, a 104.8-kb genomic region (chr2:16,452,735-16,557,612; hg38) was selected for targeted sequencing in the discovery cohort, which included 1,414 cases and 1,371 controls (Table S1). Genomic DNA was extracted from peripheral blood, sheared to approximately 250 bp and used for library construction with the SureSelect XT Target Enrichment System (Agilent, CA, USA). The Illumina HiSeq 2000 platform (PE150) was used for sequencing to achieve an average coverage depth of >30×.

Bcl2fastq was used to demultiplex the raw sequencing data, and Fastx to perform quality trimming. High-quality reads were aligned to the hg38 reference genome with Burrows-Wheeler Aligner (BWA).^35^ Variant calling was completed using GATK (v3.5)^36^ and Varscan (v2.3.9).^37^ Variants were filtered on the basis of sequencing depth (>20×) and quality score (>30) in >80% of individuals. SNPs that deviated from Hardy-Weinberg equilibrium (HWE; *P* < 0.001 in controls) or had a minor allele frequency (MAF) < 0.01 were excluded. Association analysis was performed with PLINK (v1.07)^38^ using logistic regression under an additive model, with adjustment for sex and age.

A total of 84 suggestive variants (P < 5 × 10^−3^) were further validated in an independent replication cohort comprising 1,023 cases and 1,020 controls (Tables S1 and S2). Inclusion criteria were identical to those described above. Genotyping was performed with the SNaPshot assay (Genesky Biotechnologies, Shanghai, China) and analyzed on an ABI 3130XL Gene Analyzer (Applied Biosystems, CA, USA). Quality control excluded samples with call rates <95% and SNPs with call rates <95% or deviation from HWE (P < 0.001). Association testing was performed with PLINK (v1.9).

Meta-analysis of the discovery and replication phases was performed with METAL using an inverse-variance-weighted fixed-effects model. Before meta-analysis, stringent data quality control was applied. Short insertions and deletions (indels) were excluded from the pooled meta-analysis because of allele-format incompatibilities with the METAL algorithm. The associations of these indel variants were therefore evaluated independently in the respective cohorts. Heterogeneity was assessed with Cochran’s Q test and I^2^ statistics.

### Cell culture

The human embryonic stem cell line H9 (hESC-H9) was provided by the CAS Center for Excellence in Molecular Cell Science, Shanghai Institute of Biochemistry and Cell Biology, University of Chinese Academy of Sciences (Shanghai, China), and maintained in mTeSR™1 (STEMCELL Technologies, 85850, Vancouver, Canada) on plates coated with Corning Matrigel matrix (Corning, 354277, NY, USA). The medium was changed every other day. Confluent hESCs were passaged with ReLeSR (STEMCELL Technologies, 100-0483) and cryopreserved in CryoStor® CS10 (STEMCELL Technologies, 100-1061).

The human embryonic palate mesenchymal (HEPM) cell line was obtained from the American Type Culture Collection (USA) and maintained in MEM (Gibco, 11095080, MA, USA) supplemented with 10% FBS (Gibco, 10099141) and 1% Penicillin-Streptomycin solution (P-S) (Gibco, 15140122). The human embryonic kidney 293T (HEK293T) cell line was provided by the China Type Culture Collection, Wuhan University (Wuhan, China), and maintained in DMEM (Gibco, 11995) supplemented with 10% FBS and 1% P-S. The human immortalized oral epithelial cell (HIOEC) line (a kind gift from Professor Huan Liu) was cultured in Keratinocyte SFM (Gibco, 17005042). These cells were cryopreserved in serum containing 10% DMSO (Merck, D8418, Darmstadt, Germany). Unless otherwise stated, all cells were passaged with Trypsin-EDTA (Gibco, 25200056), cryopreserved in complete medium containing 10% DMSO (Merck, D8418), and cultured with medium changes every other day.

### Differentiation of hESC-derived cNCCs

hESC-derived cNCCs were generated according to well-established protocols.^39–42^ hESCs were maintained in mTeSR™1 on Corning Matrigel matrix, and colonies were typically passaged every 4-7 days. Differentiation into cNCCs was induced with the STEMdiff Neural Crest Differentiation Kit (STEMCELL Technologies, 08610) according to the manufacturer’s instructions. In Brief, Accutase (Thermo Fisher Scientific, 00-4555-56, MA, USA) was used to dissociate hESCs into single cells. After being resuspended in the supplied medium containing 10 μM Y-27632 (Cell Signaling Technology, 13624S, MA, USA), cells were seeded at 2 × 10^5^ cells/cm^2^ onto Matrigel-coated plates. Culturing was performed by changing the medium daily without Y-27632. On day 6, cells were dissociated into single cells with Accutase and collected as passage 1 cNCCs (cNCC P1).

To induce osteogenic differentiation, cNCCs were treated with 50 μg/mL L-ascorbic acid (Merck, A4544), 100 nM dexamethasone (Merck, D4902), and 10 mM β-glycerophosphate (Merck, G9422). Cells were harvested after 7 days for reverse transcription quantitative PCR (RT-qPCR), western blotting (WB), and alkaline phosphatase (ALP) staining, and after 21 days for Alizarin Red S (ARS) staining.

### Plasmid constructs

The 1-kb sequence upstream of the *MYCN* transcription start site (promoter) and all candidate elements within the 2p24.2 region were cloned from genomic DNA extracted from HEK293T cells with the FastPure Cell/Tissue DNA Mini Extraction Kit (Vazyme, DC102, Nanjing, China). The *MYCN* promoter fragment was cloned into pGL6-TA (Beyotime, D2105, Shanghai, China) to generate pGL6-TA-Pro. Candidate element products were validated by Sanger sequencing, and genotypes at each SNP site were compared between the risk and non-risk alleles. The Mut Express II Rapid Mutagenesis Kit V2 (Vazyme, C214) was employed in Site-directed mutagenesis to generate constructs carrying either the non-risk or risk allele. After validation, these fragments were cloned into pGL6-TA-Pro for *in vitro* luciferase assays. Enh-*MYCN* fragments carrying the non-risk or risk allele were then subcloned into the cFOS-GFP reporter construct (a kind gift from professor Huan Liu), for the *in vivo* zebrafish enhancer assay. Primers were synthesized by Sangon (Shanghai, China) and are listed in Table S7.

Coding sequences of candidate TFs (listed in Table S6) were obtained from NCBI. These coding sequences were cloned into the pCMV-3×Flag plasmid (MiaolingBio, P55018, Wuhan, China) for overexpression assays during functional TF screening. The coding sequence of human *FOXP2* was also subcloned into pCMV-MCS (MiaolingBio, P51095) linked to an EGFP sequence for fluorescence recovery after photobleaching (FRAP). The intrinsically disordered region (IDR) sequence of human *FOXP2* was then obtained from NCBI and cloned into pCMV-mCherry-MCS-3 ×FLAG (MiaolingBio, P58652) for rescue experiments. All plasmids used in the overexpression, FRAP, and rescue experiments were synthesized by Miaoling Bio (Wuhan, China).

### Dual-luciferase assay

Each reporter construct was transfected with the Renilla luciferase plasmid (Beyotime, C2760) in three biological replicates. Transfection into HEPM and HIOEC cells (10^6^ cells per transfection) was performed with Lipo8000 (Beyotime, C0533) according to the manufacturer’s standard protocol. Luciferase activity was measured with the Dual-Luciferase Assay Kit (Beyotime, RG029) according to the manufacturer’s instructions using a GloMax® 20/20 Luminometer (Promega, Madison, WI, USA) at 48 h after transfection. The ratio of firefly to Renilla luciferase activity was calculated as relative luciferase activity. Three measurements were obtained from the lysates of each transfection group.

### Chromatin conformation capture (3C) and genotyping of 3C products

Primers for 3C-PCR are listed in Table S7 and were synthesized by Sangon. The 3C-PCR assay was performed according to the manufacturer’s instructions for the Chromosome Conformation Capture (3C) Kit (BersinBio, Bes5006, Guangzhou, China), with at least two replicates. Briefly, approximately 10^7^ cells were crosslinked in 1% formaldehyde and then quenched with 1.375 M glycine, followed by restriction digestion. Pellets were resuspended in 50 μL of 1× restriction enzyme buffer, lysed with 0.2% SDS, and then treated with 1.2% Triton X-100. Lysates were digested with EcoRI (NEB, R3101, MA, USA) and ligation was then performed in 1× ligation buffer at 16 °C for 2 h. The phenol/chloroform extraction was employed to purify the ligation products. The purified ligation product served as the template for 3C-PCR using a high-fidelity enzyme (Vazyme, PK511) and primers anchored near the EcoRI sites flanking Enh-*MYCN* and the *MYCN* or *DDX1* transcription start sites. The gel electrophoresis was used to separate 3C-PCR products, and bands of the target size were recovered, purified, and sequenced. Sanger sequencing was performed by Sangon.

### Generation of Enh-*MYCN* knockout hESCs

Guide RNAs (gRNAs) flanking the Enh-*MYCN* were designed computationally with the CRISPRscan algorithm. All gRNA sequences used in this study are documented in Table S7. The complete gene-editing workflow, from vector construction to delivery of verified monoclonal cell lines, was performed by Ubigene (Guangzhou, China).

Targeted genomic deletion in hESCs was achieved using a Cas9 ribonucleoprotein (RNP) delivery strategy. Briefly, a pair of synthetic single-guide RNAs (sgRNAs) was co-delivered with Cas9 protein to induce a double-strand break and excise the intervening DNA fragment. For each electroporation reaction, 3 × 10^5^ hESCs in exponential growth phase were harvested and pelleted. The cell pellet was suspended in electroporation buffer and then mixed thoroughly with a preformed RNP complex containing 50 pmol sgRNA duplex and 3 μg EZ-editor™ Cas9-EGFP protein (Ubigene, YK-Cas9-50). The 10 μL cell-RNP suspension was electroporated with the Neon™ Transfection System (Thermo Fisher Scientific, MPK10096) with the following conditions: 1320 V, 30 ms, and 2 pulses.

After electroporation, cells were cultured for 48 h. To assess editing efficiency, a fraction of the transfected cell pool was collected. Genomic DNA was extracted with a Genomic DNA Extraction Kit (TIANGEN, DP304, Beijing, China). The target locus was amplified with primers flanking the intended cleavage sites by PCR, and Sanger sequencing was performed on the purified PCR products.

The remaining successfully edited cells were dissociated into single cells and seeded into 96-well plates by limiting dilution to allow monoclonal colony formation. After approximately 2 weeks, visible colonies were replicated into two 96-well plates. Genomic DNA was extracted from one plate for screening, whereas the other plate was maintained as a master plate for clone expansion. Clones carrying the desired biallelic deletion were identified by PCR amplification with primers flanking the target region, followed by confirmation by Sanger sequencing. Verified knockout-positive clones were then thawed, expanded, and cryopreserved for subsequent experiments.

### Establishment of isogenic hESC rs4263114 mutant lines

To generate isogenic hESC lines carrying the homozygous T/T or G/G genotype, the CRISPR/Cas9-mediated knock-in strategy was used in the parental wild-type hESC line, which harbored the heterozygous T/G genotype. The complete gene-editing procedure, from vector construction to delivery of verified monoclonal cell lines, was performed by Ubigene.

This strategy used an all-in-one plasmid and a single-stranded oligodeoxynucleotide (ssODN) repair template. Briefly, a single-guide RNA (sgRNA) was designed to target the genomic region containing the SNP of interest. The sgRNA was cloned into the CRISPR-U™ expression plasmid (Ubigene), which co-expresses Cas9 nuclease and a puromycin-resistance gene for selection. For each intended point mutation (T→G or G→T), a corresponding 100-bp ssODN repair template was synthesized. Each ssODN carried the desired nucleotide substitution and silent mutations within the PAM sequence to prevent Cas9 re-cleavage after successful homology-directed repair (HDR). All primers and oligonucleotides used in this study are detailed in Table S7.

For electroporation, approximately 1 × 10^6^ hESCs were co-transfected with 30 μg of the CRISPR-U™ plasmid and the corresponding ssODN donor utilizing the Neon™ Transfection System (Thermo Fisher Scientific) under conditions of 1320 V, 30 ms, and 2 pulses. At 48 h after transfection, puromycin selection was used to enrich for successfully transfected cells. Then, to isolate monoclonal colonies, surviving cells were dissociated and seeded into 96-well plates with limiting dilution.

After 2-4 weeks, individual clones were screened by PCR amplification and Sanger sequencing to identify clones carrying the precise homozygous T/T or G/G substitution. Verified positive cell lines were then expanded for subsequent experiments.

### Reverse transcription quantitative PCR analysis

The FastPure Total RNA Isolation Kit (Vazyme, RC112) and All-in-one RT SuperMix for qPCR (Vazyme, R333) were used in total RNA extraction and reverse transcription separately. RT-qPCR was then conducted on a CFX96 Real-Time PCR Detection System (Bio-Rad, CA, USA) according to the manufacturer’s guidelines for ChamQ Universal SYBR qPCR Master Mix (Vazyme, Q711). *GAPDH* was used as an intrinsic control. Primers are documented in Table S7 and were synthesized by Sangon. Relative expression was calculated with the 2^−ΔΔCt^ method.

### Western blotting

Cell lysis buffer for Western blotting (Beyotime, P0013) supplemented with 1× PMSF (Beyotime, ST506) was used to extract proteins from cultured cells. Lysates were sonicated and centrifuged at 4 °C and the supernatants were collected into fresh tubes. Protein concentration was tested using the Enhanced BCA Protein Assay Kit (Beyotime, P0010). Protein samples were mixed with SDS-PAGE Loading Buffer (Biosharp, BL1012A, Hefei, China) and denatured at 95 °C for 5 min.

For SDS-PAGE, proteins were separated on 10% gels with SWE Rapid High Resolution Running Buffer (Servicebio, G2081-15, Wuhan, China) and then transferred onto PVDF Western Blotting Membranes (Roche, 03010040001, Basel, Switzerland) using Ice-Bath Free Fast Transfer Buffer (Servicebio, G2028-15). Membranes were blocked with QuickBlock™ Western Blocking Solution (Beyotime, P0252) and incubated with primary antibodies at 4 °C for 12-16 h. After three washes with 1× TBST (Solarbio, T1082, Beijing, China), membranes were incubated with the appropriate HRP-conjugated secondary antibodies. Antibodies used in this study are detailed in Table S8.

### ALP and ARS staining

For ALP staining, 7-day osteogenically induced cells were fixed in 4% paraformaldehyde (PFA) (Servicebio, P1110) and washed with 1× phosphate-buffered saline (PBS) (Gibco, 10010023). Staining was performed using the BCIP/NBT Alkaline Phosphatase Color Development Kit (Beyotime, C3206).

For ARS staining, cells were cultured under osteogenic conditions for 21 days and stained with Alizarin Red S Solution (OriCell, ALIR-10001, Suzhou, China). Stained cells were imaged using a stereo microscope (Olympus SZX7, Japan) and an inverted microscope (Olympus CKX53).

### Cell proliferation and apoptosis assays

For proliferation assays, the logarithmic growth phase cNCCs were seeded into 96-well plates at 5 × 10³ cells per well. After cell attachment, 10 μL of Cell Counting Kit-8 (CCK-8; Biosharp, BS350B) was added to each well and the plates were incubated at 37 °C for 1 h at 0, 24, 48, and 72 h. The absorbance was measured at a wavelength of 450 nm using a microplate reader (BioTek Synergy LX, VT, USA).

The Annexin V-FITC Kit (Absin, abs50001, Shanghai, China) was employed to assess apoptosis in flow cytometry. In brief, cells (1 × 10^6^ per replicate) were washed with 1× PBS and resuspended in 300 μL of 1× binding buffer. After staining with 5 μL Annexin V-FITC for 15 min in the dark, cells were treated with 5 μL propidium iodide (PI) before flow-cytometric analysis on a CytoFLEX LX flow cytometer (Beckman Coulter, CA, USA). Data were analyzed using CytExpert software (Beckman Coulter).

### Jawbone-like organoid construction

Jawbone-like organoids were generated according to a previously reported three-dimensional stepwise induction protocol.^43^ Briefly, after dissociation, hESCs were seeded into Nunclon™ Sphera™ 96-well plates (Thermo Fisher Scientific, 174929) and cultured with StemFit Basic03 medium (Ajinomoto, SF031-001, Tokyo, Japan) containing 10 μM Y-27632. To promote cell aggregation, plates were centrifuged at 280 × g for 3 min. The medium was changed to 150 μL per well of StemFit Basic03 containing 10 μM SB431542 (MCE, HY-10431, NJ, USA) and 10 ng/mL BMP4 (MCE, HY-P7007) after 24 h. After an additional 24 h, cell aggregates were individually transferred to BeyoGold™ Ultra-Low Attachment 48-well plates (Beyotime, FULA-485) and maintained in 250 μL of StemFit Basic03 per well, which was supplemented with 10 μM SB431542 and 1 μM CHIR99021 (Selleck, S2924, TX, USA). Plates were then placed on a Thermo Scientific™ CO^2^-resistant shaker (Thermo Fisher Scientific, 88881104) and cultured at 120 rpm until day 5.

On day 5, the medium was refreshed with 350 μL per well of StemFit Basic03 containing the FEDB combination: 100 ng/mL FGF8b (MCE, HY-P70533), 50 nM endothelin-1 (EDN1) (MCE, HY-P0202), and 2.5 ng/mL BMP4. Aggregates were cultured in the 48-well plates on a 120-rpm shaker for a further 4 days until day 9. The mdEM aggregates obtained on day 9 were transferred to new Nunclon™ Sphera™ 96-well plates, and the medium was changed to StemFit Basic03 containing 20 ng/mL EGF (PeproTech, AF-100-15, NJ, USA), 20 ng/mL FGF2 (PeproTech, 100-18B), 50 ng/mL BMP2 (MCE, HY-P7006), 3 μM CHIR99021, and 400 nM smoothened agonist (SAG) (MCE, HY-12848).

On day 12, the medium was replaced with 200 μL per well of MSCgo Osteogenic Differentiation Medium (STEMCELL Technologies, 05465). On day 19, StemFit Basic03 was added at 200 μL per well containing the following osteogenic inducers: 10 mM β-glycerophosphate, 250 μM L-ascorbic acid, 100 nM dexamethasone, 50 ng/mL BMP2, 3 μM CHIR99021, and 400 nM SAG. On day 26, the medium was again refreshed with MSCgo Osteogenic Differentiation Medium and changed every 2 days until harvest on day 38.

### Immunofluorescence

For jawbone-like organoids, samples were fixed in 4% PFA, dehydrated in 30% sucrose (Hushi, 8969, Shanghai, China), and embedded in FSC 22 Frozen Section Compound (Leica, 3801480, USA). Sections (7 μm) were incubated with Blocking Buffer (Beyotime, P0260). Then, primary antibodies and secondary antibodies were used for incubation at 4 °C and 37 °C, separately. Nuclei were counterstained with DAPI, and images were acquired using a confocal microscope. Relative mean fluorescence intensity was quantified using ImageJ (NIH, MD, USA) from three biologically independent experiments for each group.

For immunocytochemistry, cells were fixed in 4% PFA and permeabilized with 0.3% Triton X-100 (Biofroxx, 1139, Einhausen, Germany). Cells were then incubated with Blocking Buffer at 37°C for 15 min, followed by incubation with primary and secondary antibodies. Images were acquired using a confocal microscope.

Antibodies used in this study are documented in Table S8.

### Enhancer reporter assay in zebrafish

The cFOS-GFP vector carrying each candidate fragment was co-injected with Tol2 mRNA (20 ng/μL) into one-cell-stage zebrafish embryos. All healthy embryos were screened at 72 h after injection by fluorescence microscopy in an unblinded mantablener to compare tissue-specific GFP expression patterns and assess enhancer activity during embryonic development.

To visualize LLPS of FOXP2 in vivo, the mMESSAGE mMACHINE SP6 Kit (Thermo Fisher Scientific, AM1340) was used to generate capped mRNAs encoding EGFP-FOXP2 or EGFP alone through *in vitro* transcription. pCS2+ plasmids carrying the corresponding cDNA sequences (purchased from MiaoLingBio) were used as templates. Purified mRNAs were then microinjected into wild-type single-cell zebrafish embryos. Phase-separated condensates were monitored and imaged at 2 h post-fertilization (hpf).

### TF prediction and validation

Candidate TFs were predicted using the JASPAR database. Their expression during craniofacial development, from neural crest cells to Carnegie stage 22 (NCC-CS22), was then evaluated using bulk RNA-sequencing data from human craniofacial tissues^44^ and single-cell RNA-sequencing data from mouse and human craniofacial tissues.^44^ Only TFs expressed in mesenchymal cells before CS22 in human craniofacial tissues were retained for validation (Table S6).

Overexpression constructs were transfected into hESCs (1 × 10^6^ cells per transfection) using Lipomaster 3000 Transfection Reagent (Vazyme, TL301). Cells were harvested for RT-qPCR analysis of *MYCN* and TF expression at the transcriptional level 48 h after transfection. The RT-qPCR procedure is described above. Primers are detailed in Table S7 and were synthesized by Sangon.

### Chromatin immunoprecipitation-quantitative PCR (ChIP-qPCR)

The ChIP Kit Magnetic - One Step (Abcam, ab156907, Cambridge, UK) was used in Chromatin immunoprecipitation. In brief, approximately 1 × 10^7^ cells were crosslinked with 1% formaldehyde and the reaction was quenched with glycine. Then, cells were subjected to a two-step lysis procedure: an initial incubation in hypotonic cell lysis buffer on ice to isolate nuclei, followed by nuclear lysis with SDS lysis buffer (Beyotime, P0013G).

Chromatin was sheared to 200–500 bp size fragments using a probe sonicator (Sonics Organomation VCX130, TX, USA). Sonication was performed on ice at 8% power output in pulse mode (5 s on and 5 s off) for a total of 8 min. Shearing efficiency was confirmed by agarose gel electrophoresis.

For immunoprecipitation, the sheared chromatin was incubated with 2 μg of either anti-FOXP2 antibody (Novus Biologicals, NB100-55411, CO, USA) or normal rabbit IgG (Cell Signaling Technology, 2729S). Antibody-chromatin complexes were captured with Protein A/G magnetic beads. After a series of stringent washes according to the kit protocol, the immunoprecipitated (IP) chromatin was eluted and crosslinks were reversed. DNA was then purified after proteinase K digestion.

Purified DNA from both IP and input samples was quantified by qPCR using primers designed to amplify specific genomic regions. Primer sequences are documented in Table S7 and were synthesized by Sangon. A previously validated FOXP2-binding site (chr3:194,685,617-194,686,023; hg38) served as the positive control region.^45^ A distal gene-desert region (chr2:15,948,955-15,949,053; hg38) was employed as the negative control region to assess non-specific binding.

### Fluorescence recovery after photobleaching

For FRAP assays, after transfecting with the indicated constructs, HEPM cells were imaged using an Olympus microscope with a 100× oil-immersion objective. On the basis of the initial fluorescence intensity, droplets were selected and bleached for 1 s at 50% laser power. Approximately 50-70% of the signal was bleached under these conditions. Time-lapse images were taken every 15 s for 3 min. Fluorescence intensity was quantified using ImageJ.

### Statistical analyses

All quantitative experiments, including RT-qPCR, dual-luciferase assays, immunofluorescence assays and ChIP-qPCR, were conducted with three independent biological replicates. Three technical replicates were included for each biological replicate.

The GraphPad Prism 8 was used in statistical analyses and graph generation. Data are presented as mean ± SD. *P* values and Sample size (*n*) are shown in the figures. Data normality was assessed using the Shapiro-Wilk test. After equality of variance had been confirmed by an F-test, an unpaired, two-tailed Student’s t-test was performed in comparisons between two groups. For comparisons among three or more groups, one-way or two-way analysis of variance (ANOVA) was used as appropriate, incorporating Tukey’s or Sidak’s post hoc test.

## Results

### Identification of a candidate functional variant within a regulatory element at 2p24.2

Previous GWAS studies have established rs7552 as a tag SNP for NSCLP across diverse ancestral populations.^23,46,47^ To comprehensively identify potential functional variants within this locus, we first outlined the linkage disequilibrium (LD) block linked to rs7552. Using PLINK, we defined a 104-kb genomic interval spanning chr2:16,452,735-16,557,612 (hg38) (Figure S1A).

In the discovery stage, we performed targeted sequencing using a custom Agilent SureSelect panel in a cohort comprising 1,414 NSCLP cases and 1,371 controls of Han Chinese ancestry (Table S1). Following quality control, variants with a genotype missingness greater than 5% were excluded from further analysis. Additionally, SNPs that significantly deviated from HWE (*P* < 1 × 10^−3^) or had a MAF < 0.01 were removed. A total of 292 highly qualified SNPs were advanced to the discovery analysis. We then assessed genotype-phenotype associations using logistic regression under an additive model, with adjustment for sex and age. We prioritized 84 candidate SNPs (*P* < 5 × 10^−3^) for genotyping in a replication cohort comprising 1,023 NSCLP cases and 1,020 controls of Han Chinese ancestry. In the replication stage, all 84 SNPs were successfully genotyped (Table S2), and 62 showed nominally significant association (*P* < 0.05, logistic regression), all with effect directions consistent with those in the discovery stage.

In the subsequent fixed-effects meta-analysis combining both cohorts, totaling 2,437 NSCLP cases and 2,391 controls, we identified 55 SNPs with *P* < 5 × 10^−8^ (Figure 1A and Table S2). The overall study workflow is summarized in Figure 1B.

**Figure 1.**
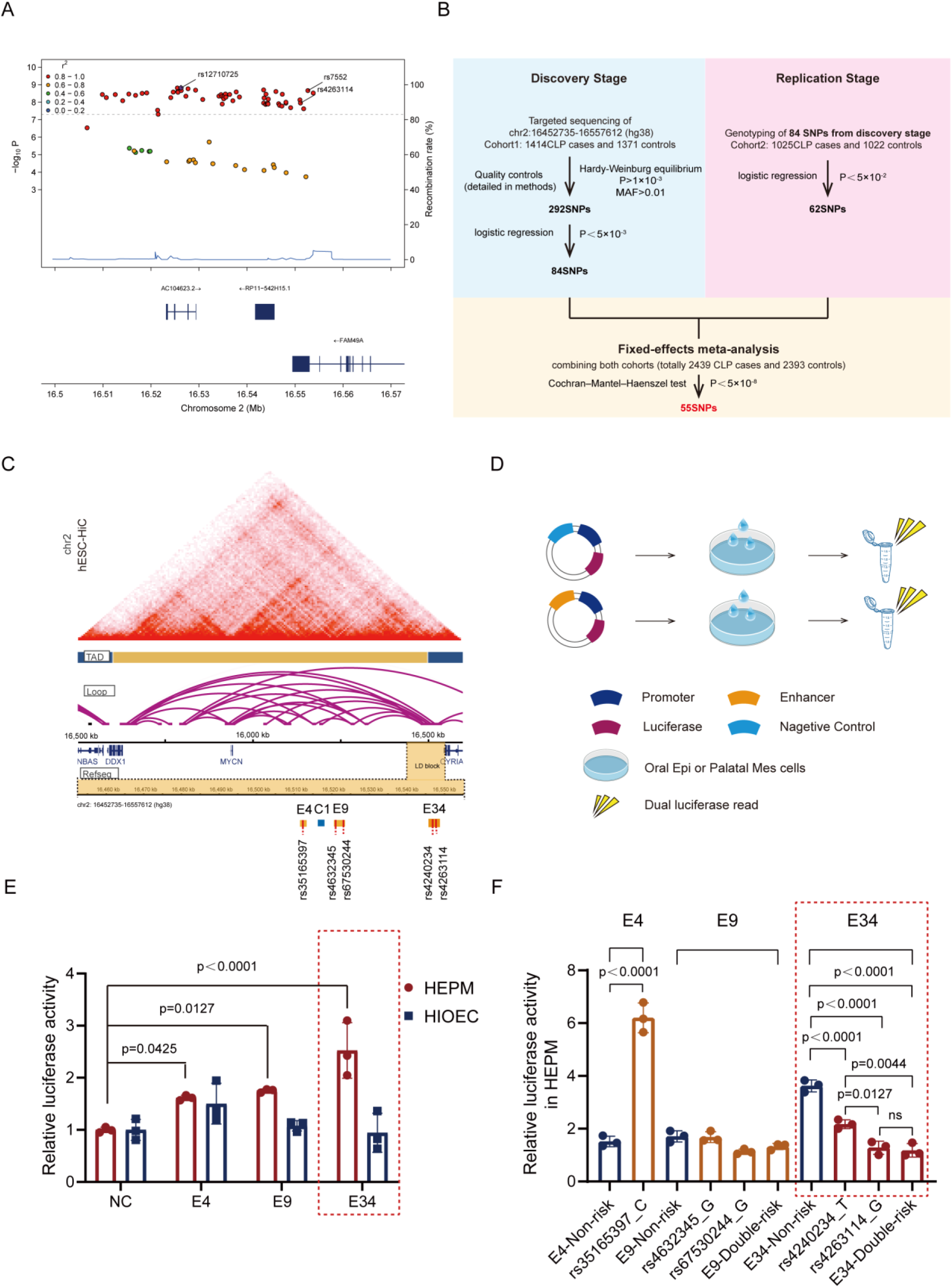
Discovery and functional evaluation of non-coding regulatory elements carrying NSCLP-risk SNPs within the 2p24.2 region. **(A)** Regional association analysis highlighting the 104-kb LD block tagged by rs7552. LocusZoom plot derived from the meta-analysis of our Chinese NSCLP cohorts. Each dot signifies a variant, with its color reflecting the LD (*r*^2^) relative to the current index SNP rs12710725 (purple), which exhibits strong correlation with the formerly identified top SNP rs7552. The functional enhancer SNP rs4263114 is also marked in the figure. **(B)** Schematic overview of the two-stage genetic screening strategy. **(C)** Spatial chromatin architecture of the 2p24.2 interval visualized by Hi-C in human embryonic stem cells (H9 line; GSE116862). The identified linkage disequilibrium (LD) block is highlighted, showing the genomic positions of the negative control fragment (C1) and three functional candidate elements (yellow), together with their respective candidate SNPs (red). **(D)** Schematic illustration of the luciferase reporter assay used to screen enhancer activity of the cloned candidate elements. **(E)** Candidate elements exhibit mesenchymal-specific enhancer activity. Luciferase reporter assays for three functional enhancers alongside the C1 control in HIOEC and HEPM cell lines. Significance is shown relative to the C1 control within each cell line. Mean ± SD (*n* = 3). Two-way ANOVA (Dunnett’s post-hoc test). **(F)** Candidate SNPs drive allele-specific enhancer activity. Luciferase reporter assays comparing constructs harboring either the non-risk or risk alleles in HEPM cells. Mean ± SD (*n* = 3). One-way ANOVA (Tukey’s post-hoc test).

By integrating our fine-mapping results with epigenomic datasets from FaceBase, three of the 55 candidate SNPs were located within previously reported enhancer regions^48^ (Figure S1B). To systematically screen for functional elements, we grouped the candidate SNPs into clusters based on LD (*r*^2^< 0.8) and physical proximity (<1 kb). We then defined 37 potential enhancer fragments (E1-E37) by extending approximately 500 bp upstream and downstream of the candidate SNPs in each cluster (Figure 1B and Table S3). Additionally, we selected a genomic fragment (C1) within this topologically associating domain (TAD) as a negative control because it lacked enhancer-associated chromatin signatures in human embryonic cranial tissues (Figure 1C, Figure S1B, and Table S3).^49^

We individually cloned all 38 elements upstream of the *MYCN* promoter into a firefly luciferase reporter vector, each of which was co-transfected with a *Renilla* luciferase vector as a transfection control into HEPM cells, and luciferase activity was assessed after 48 h (Figure 1D). Whereas the negative control C1 showed minimal background activity, fragments E4 and E9 exhibited approximately twofold higher activity, and E34 displayed a robust threefold increase relative to C1 (Figure 1E and Figure S1C). By contrast, these three fragments showed no enhancer activity in oral epithelial HIOEC cells, indicating tissue specificity (Figure 1E). Importantly, the E36 fragment harboring rs7552 did not show significant enhancer activity (Figure S1C), supporting the hypothesis that rs7552 functions primarily as a genetic marker rather than a causal variant.

We next investigated whether these functional SNPs exert allele-dependent effects on enhancer activity. Using site-directed mutagenesis, we introduced risk alleles into the enhancer constructs and repeated the luciferase assays in HEPM cells (Figure 1F). For enhancers E9 and E34, each harboring two SNPs, we generated vectors carrying either single mutant SNPs or combined double mutations. The activity of E9 was unaffected by the risk alleles of rs4632345 and rs67530244, suggesting that these SNPs are unlikely to be functional drivers (Figure 1F). For E4, the risk allele of rs35165397 increased enhancer activity nearly fourfold, a pattern inconsistent with the expected loss-of-function mechanism (Figure 1F). By contrast, within E34, enhancer activity was reduced by 40% and 65% when rs4240234 and rs4263114 carried the risk allele, respectively, compared to the non-risk allele. Notably, the reduction in E34 activity observed with both risk alleles was nearly identical to that observed with rs4263114 alone (Figure 1F). These results demonstrate that rs4240234 may act as a co-contributor with rs4263114, whereas rs4263114 exerts the dominant effect on E34 enhancer activity and is therefore the more likely functional SNP.

Haplotype analysis of rs4240234 and rs4263114 within E34 is presented in Table S4. The haplotype carrying both risk alleles showed a significantly stronger genetic association with NSCLP. Based on the combined functional and genetic evidence, we selected E34 for further investigation.

### E34 physically interacts with the *MYCN* promoter to regulate its expression and cNCC differentiation

To determine the endogenous target gene of the E34 enhancer, we performed 3C assays in hESC-derived cNCCs, which were validated by specific markers (Figures S2A-S2D).^42^ The 3C analysis revealed a specific PCR product linking the E34 region (EcoRI restriction fragments) to the *MYCN* promoter, confirming their physical proximity and the assembly of a long-range chromatin loop (Figures 2A-C). In contrast, no interplay was observed between E34 and the promoter of *DDX1*, the other candidate gene at 2p24.2 (Figures S2E and S2F), suggesting that E34 specifically targets *MYCN*.

**Figure 2.**
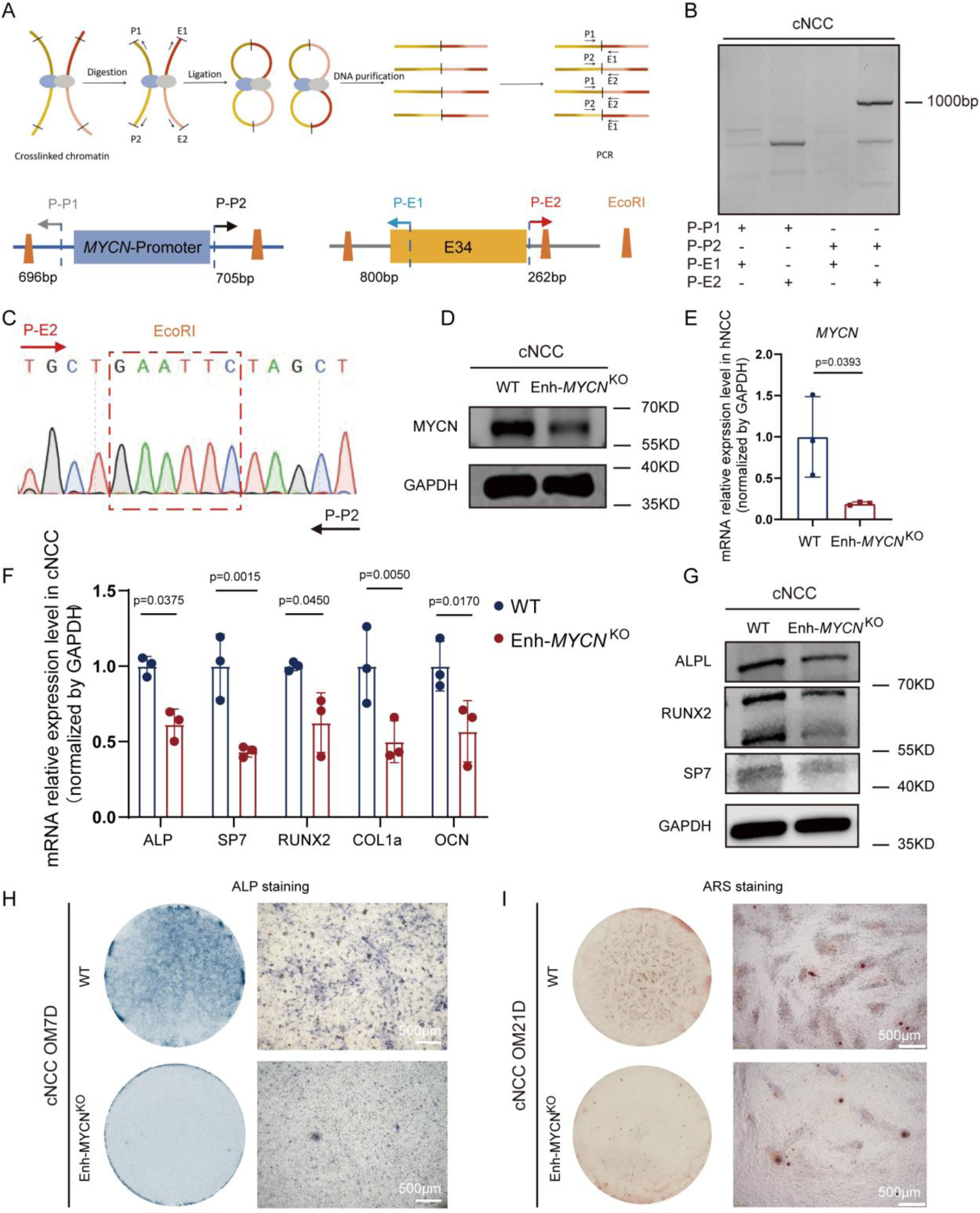
E34 physically interacts with the *MYCN* promoter to modulate its expression and govern cNCC osteogenic differentiation. **(A-C)** E34 forms a distal interplay with the *MYCN* promoter. (A) Schematic of the chromatin conformation capture (3C) assay design. EcoRI restriction sites flanking E34 and the *MYCN* promoter are indicated by orange trapezoids. Primer positions relative to the EcoRI sites are denoted by colored arrows (P-P1, 696 bp; P-P2, 705 bp; P-E1, 800 bp; P-E2, 262 bp upstream of the respective restriction sites). (B) Agarose gel electrophoresis of the 3C-PCR product (967 bp) amplified using the P-P2 and P-E2 primer pair in cNCCs. (C) Sanger sequencing chromatogram of the 3C-PCR product, confirming the presence of the ligated EcoRI restriction site (red dashed box). **(D and E)** Knockout of E34, hereafter designated Enhancer-*MYCN* (Enh-*MYCN*), downregulates *MYCN* expression. Validation of *MYCN* loss at both protein (D) and transcript levels (E) in cNCCs lacking the Enh-*MYCN* compared to WT cells. Data normalized to *GAPDH*. Mean ± SD (*n* = 3). Unpaired two-tailed Student’s t-test. **(F and G)** Enh-*MYCN* deficiency impairs the expression of osteogenic differentiation markers. (F) RT-qPCR and (G) Western blot analyses of key osteogenic markers (*ALP*, *SP7*, *RUNX2*, *COL1A1*, and *OCN*) in WT and Enh-*MYCN* ^KO^ cNCCs after 7-day differentiation. Data normalized to *GAPDH*. Mean ± SD (*n* = 3). Two-way ANOVA (Sidak’s post-hoc test). **(H and I)** Enh-*MYCN* deletion suppresses cNCC osteogenic differentiation and mineralization. Representative images of (H) ALP staining at day 7 and (I) ARS staining after 21-day osteogenic induction. Scale bars, 500 μm.

To functionally validate the regulatory role of E34, we generated an isogenic knockout cell line (hESC^E34KO^) using CRISPR/Cas9-mediated deletion (Figure S3A). Sanger sequencing confirmed precise excision of the enhancer element, and pluripotency markers remained unchanged (Figures S3B and S3C). We then differentiated both wild-type (hESC^WT^) and knockout (hESC^E34KO^) lines into cNCCs. RT-qPCR and WB analyses showed a significant reduction in *MYCN* expression in cNCC^E34KO^ cells compared with cNCC^WT^ cells across both the transcriptional and translational processes (Figures 2D and 2E). Importantly, *DDX1* expression remained unchanged in the knockout cells (Figure S3D). These results confirmed that E34 specifically regulates *MYCN* and is hereafter designated Enhancer-*MYCN* (Enh-*MYCN*).

We next investigated whether reduced *MYCN* expression after Enh-*MYCN* knockout was sufficient to impair the differentiation capacity of cNCCs *in vitro*. Whereas cell proliferation and apoptosis did not differ between genotypes (Figures S3E and S3F), the knockout cells exhibited a marked defect in osteogenic differentiation. This defect was evidenced by significant downregulation of the osteogenic markers *ALP* [MIM: 171760], *SP7* [MIM: 606633], *RUNX2* [MIM: 600211], *COL1A1* [MIM: 120150], and *OCN* [MIM: 112260] (Figures 2F and 2G), coupled with diminished matrix mineralization, as visualized by ALP and ARS assays (Figures 2H and 2I). These findings confirm that Enh-*MYCN* is required to maintain *MYCN* expression at levels necessary for cNCC differentiation.

### Validation of Enh-*MYCN* activity and tissue specificity in human jawbone-like organoids and zebrafish

To investigate the function of Enh-*MYCN* in a complex tissue context, we generated human jawbone-like organoids from hESC^WT^ and hESC^Enh-*MYCN*^ *^KO^* lines using an established protocol.^43^ We confirmed the stage-specific developmental trajectory of these organoids (Figures S4A-S4C). After the same 38-day induction period, organoids derived from hESC^Enh-^*^MYCN^ ^KO^* cells were markedly smaller, showing an approximately 30% reduction in mean radius compared with controls (Figures S4D and S4E). Immunofluorescence analysis identified a striking downregulation in the expression of key osteogenic differentiation markers (SP7, COL1A1, ALP, and OCN) in the knockout group relative to controls (Figures 3A and 3B). Furthermore, markers of mature osteocytes, including PDPN (*PDPN* [MIM: 608863]) and SOST (*SOST* [MIM: 605740]), were consistently downregulated (Figures 3C and 3D). These data indicate that deletion of Enh-*MYCN* severely impairs mineralization and bone formation in a 3D craniofacial model, consistent with our *in vitro* findings.

**Figure 3.**
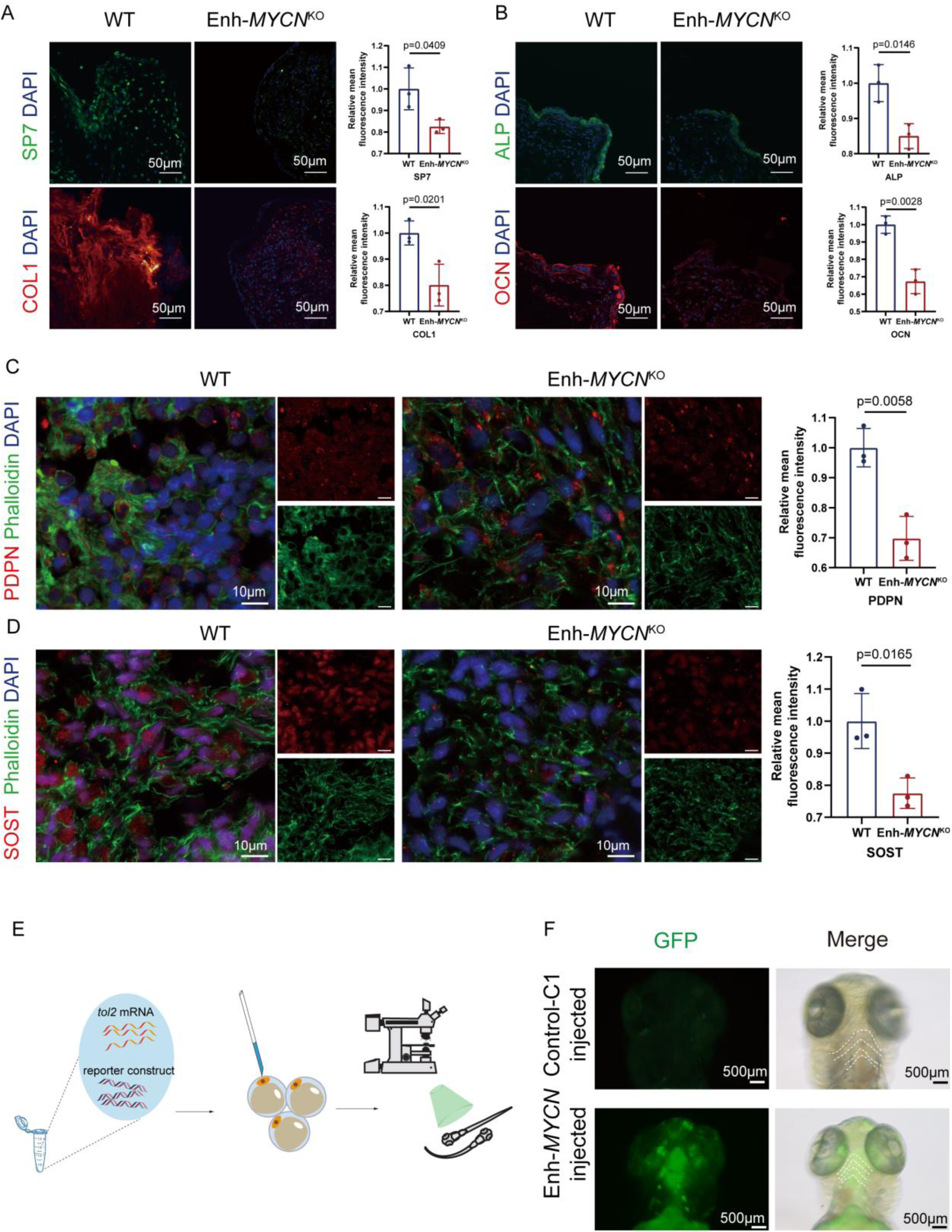
Enh-*MYCN* shows *in vivo* tissue-specific activity and is required for osteogenic differentiation in 3D jawbone-like organoids. **(A and B)** Deletion of Enh-*MYCN* impairs the expression of early osteogenic markers. Representative immunofluorescence images and corresponding quantification of (A) SP7 and type I collagen (COL1), and (B) ALP and OCN in wild-type and Enh-*MYCN* knockout human jawbone-like organoids at day 38 (d38) of differentiation. Nuclear counterstain: DAPI. Scale bars, 50 μm. Mean ± SD (*n* = 3). Unpaired two-tailed Student’s t-test. **(C and D)** Enh-*MYCN* deficiency downregulates mature osteocyte markers. Representative immunofluorescence images and quantification of (C) PDPN and (D) SOST, co-stained with F-actin (phalloidin), in d38 mandibular organoids. Nuclear counterstain: DAPI. Scale bars, 10 μm. Mean ± SD (*n* = 3). Unpaired two-tailed Student’s t-test. **(E and F)** Enh-*MYCN* drives robust and specific expression in cranial neural crest-derived tissues *in vivo*. (E) Schematic illustration of the zebrafish microinjection reporter assay. (F) Representative ventral views of wild-type zebrafish embryos at 72 hours post-fertilization (hpf) injected with GFP reporter constructs containing either Enh-*MYCN* or the negative control sequence (C1). Scale bars, 500 μm.

To further define the *in vivo* activity and tissue specificity of Enh-*MYCN*, we used a zebrafish reporter model. Enh-*MYCN* and the negative control fragment C1 were cloned into the pTg-cFOS-GFP-Tol2 backbone and microinjected into single-cell zygotes (*n* > 200). GFP expression was assessed in healthy F0 embryos at 72 hpf (Figures 3E and 3F). Embryos injected with Enh-*MYCN* showed robust GFP expression predominantly in the pharyngeal arches, the conserved embryonic primordia of the vertebrate craniofacial complex (Figure 3F). By contrast, GFP signal was absent from the epithelium and was rarely detected in embryos injected with the C1 control (Figure 3F). These results provide compelling *in vivo* evidence that Enh-*MYCN* has intrinsic enhancer activity specifically in cranial neural crest-derived tissues.

### Risk allele of rs4263114 attenuates Enh-*MYCN* function

To detect the impact of the rs4263114 risk allele on enhancer function *in vivo*, we generated zebrafish reporter constructs harboring either the non-risk or risk allele. Comparative microinjection analysis showed that embryos injected with the risk-allele construct had significantly weaker GFP in the pharyngeal arches (Figure 4A), with a substantially lower proportion of GFP-positive embryos than the non-risk group (Figures 4B, S4H, and Table S5). These *in vivo* findings are consistent with our dual-luciferase results (Figures 2C and 2D), demonstrating that the risk allele compromises the regulatory activity of Enh-*MYCN* in a physiological context.

**Figure 4.**
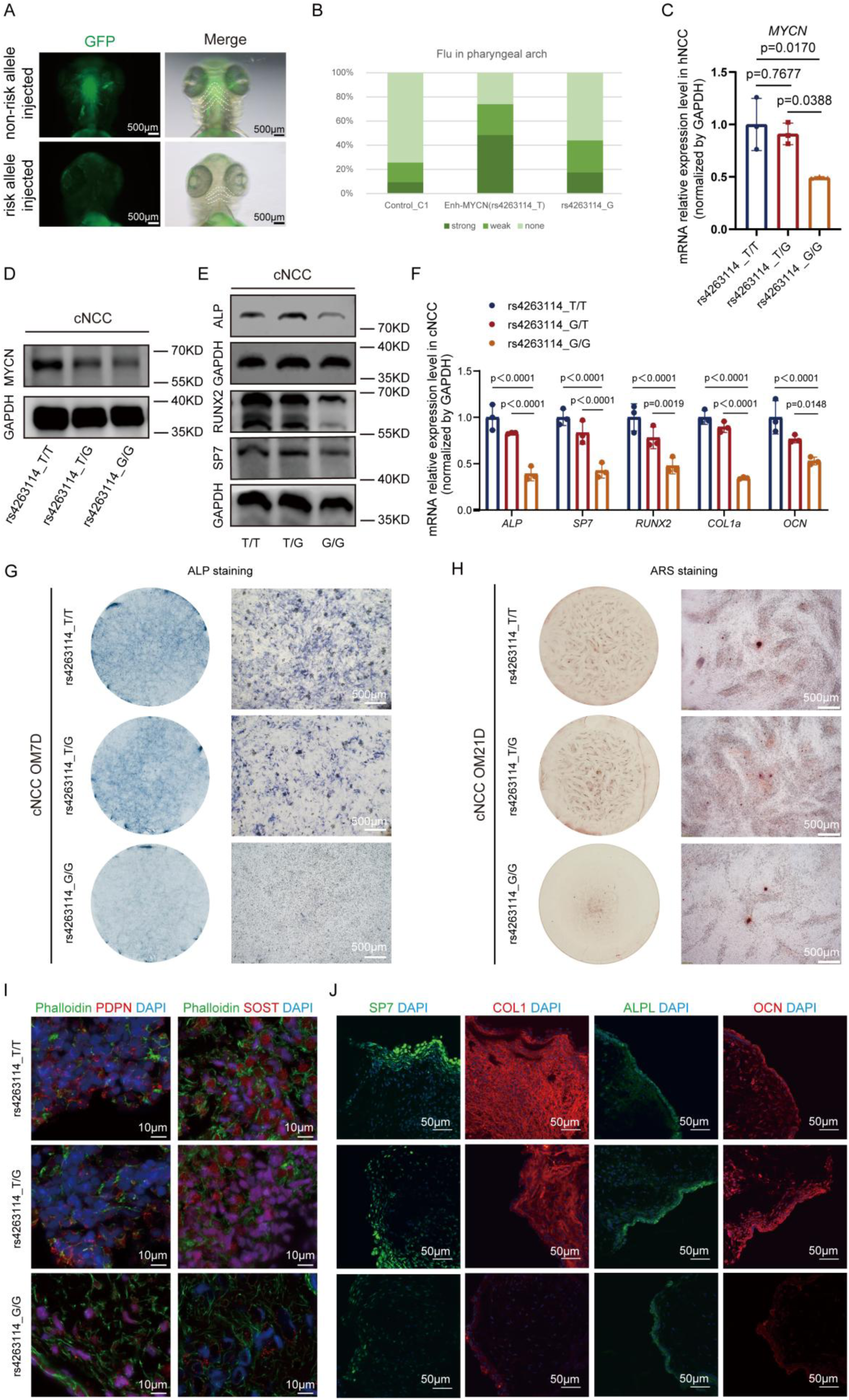
The rs4263114 risk allele dampens Enh-*MYCN* activity and impairs cNCC osteogenic differentiation. **(A and B)** The risk allele diminishes *in vivo* enhancer activity in zebrafish. (A) Representative ventral photos of zebrafish embryos at 72 hpf injected with GFP reporter constructs harboring either the Enh-*MYCN* non-risk sequence (rs4263114_T) or risk sequence (rs4263114_G). Scale bar, 500 μm. (B) One hundred percent stacked column chart exhibiting the relative percentages of zebrafish embryos with strong, weak, or no GFP fluorescence in the pharyngeal arches across all experimental groups. The total numbers of evaluated embryos injected with the C1 control, the non-risk Enh-*MYCN* allele, and the risk Enh-*MYCN* allele were 578, 593, and 556, respectively. **(C and D)** The homozygous risk allele downregulates *MYCN* expression. (C) RT-qPCR and (D) western blot were performed to evaluate mRNA and protein levels, respectively, demonstrating a significant decline in *MYCN* expression within cNCCs harboring the G/G genotype. Data normalized to *GAPDH*. Mean ± SD (*n* = 3). One-way ANOVA (Tukey’s post-hoc test). **(E and F)** The rs4263114 risk allele impairs the expression of osteogenic markers in cNCCs. (E) mRNA and (F) protein quantification of key osteogenic markers (ALP, SP7, RUNX2, COL1A1, and OCN) in T/T, T/G, and G/G cNCCs after 7 days of osteogenic induction. Data normalized to *GAPDH*. Mean ± SD (*n* = 3). Two-way ANOVA (Tukey’s post-hoc test). **(G and H)** Osteogenic differentiation and mineralization are severely impaired in risk-allele cNCCs. Representative images of (G) ALP staining at day 7 and (H) ARS staining at day 21 of osteogenic differentiation across the three genotypes. Scale bars, 500 μm. **(I and J)** The risk allele disrupts osteogenic maturation in 3D organoids. Representative immunofluorescence images of (I) mature osteocyte markers (PDPN and SOST, F-actin counterstained by phalloidin; scale bar, 10 μm) and (J) osteogenic markers (SP7, COL1, ALP, and OCN; scale bar, 50 μm) in d38 jawbone-like organoids derived from the isogenic lines. Nuclei were counterstained with DAPI.

To establish the functional causality of rs4263114, CRISPR/Cas9-mediated HDR was utilized to generate isogenic hESC lines that differed only at the rs4263114 locus. We successfully established three genotypes on the same genetic background: homozygous non-risk (T/T), heterozygous (T/G), and homozygous risk (G/G) (Figure S5A). Pluripotency marker expression remained comparable across all three lines (Figure S5B). After differentiation into cNCCs, we quantified *MYCN* expression across the three genotypes. Compared with the T/T and T/G groups, a significant reduction of *MYCN* specifically in homozygous risk (G/G) cNCCs was investigated by RT-qPCR and western blot analyses (Figures 4C and 4D). Consistent with the knockout model, *DDX1* expression remained stable, and no significant differences in cell proliferation or apoptosis were observed among the genotypes (Figures S5C-S5E).

However, cNCCs harboring the homozygous risk allele showed impaired osteogenic potential, as indicated by reduced expression of mineralization markers and diminished mineralized nodule formation compared with the non-risk groups (Figures 4E-4H). To validate these findings in a complex 3D tissue context, we generated jawbone-like organoids from the isogenic lines (Figures S4D, S4F, and S4G). Organoids derived from the homozygous risk genotype showed impaired differentiation toward mature osteocytes and osteoblasts, with reduced expression of the major bone matrix protein COL1A1 and the non-collagenous protein OCN (Figures 4I, 4J, and S5F). Collectively, these data demonstrate that the rs4263114 risk allele functionally disrupts Enh-*MYCN* activity, leading to reduced *MYCN* expression and defective cNCC differentiation.

### The rs4263114 risk allele disrupts FOXP2 binding to impair *MYCN* expression

To detect the molecular mechanism by which rs4263114 modulates the regulatory interaction between Enh-*MYCN* and the *MYCN* promoter, we utilized the JASPAR database to predict TFs whose binding affinities might be affected by the alleles of this functional SNP (Table S6). To validate these candidates, we individually overexpressed each predicted TF in cNCCs and monitored *MYCN* expression (Figures 5A, 5B, and S6A-S6E). Among the candidates, FOXP2, whose binding affinity for the risk allele was predicted to be lower than that for the non-risk allele, elicited the most significant upregulation of *MYCN* (Figures 5A-5C). Consistent with this finding, previous studies have proved the widespread expression of FOXP2 in the craniofacial region during Carnegie stages 13-17 (CS13-CS17), ^45,50^ supporting its role as a potential transcriptional activator. In parallel, we also examined TFs predicted to bind another linked SNP in Enh-*MYCN*, rs4240234. However, none of the candidate TFs was experimentally validated in a manner consistent with the prediction (Figures S6F-S6K and Table S6).

**Figure 5.**
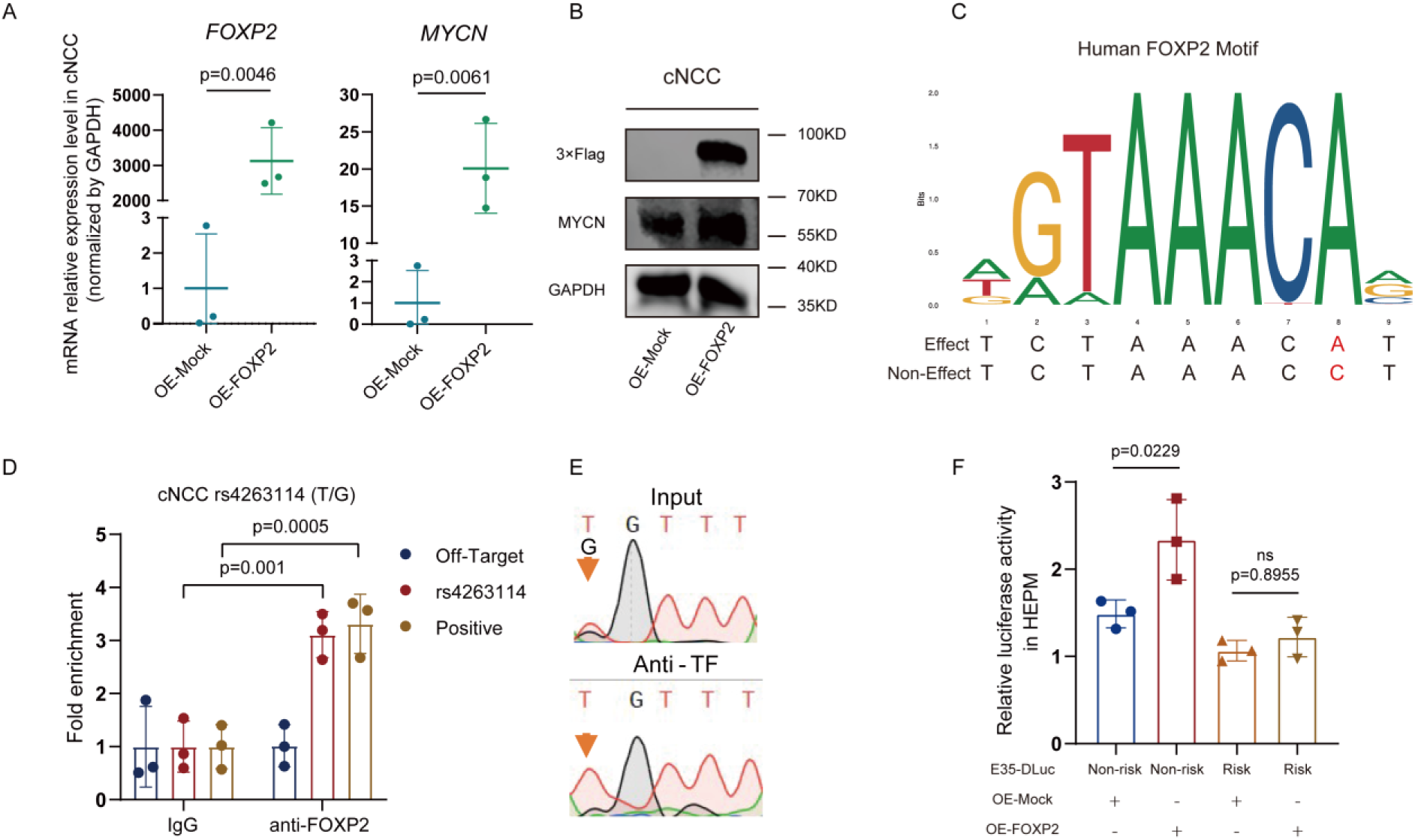
The rs4263114 risk allele compromises FOXP2 binding affinity and reduces *MYCN* expression. **(A and B)** FOXP2 overexpression upregulates *MYCN*. (A) Quantification of mRNA (RT-qPCR) and (B) protein levels (western blot) for *FOXP2* and *MYCN* expression in cNCCs transfected with a FOXP2 overexpression vector (OE-FOXP2) or an empty vector control (OE-Mock). Data normalized to *GAPDH*. Mean ± SD (*n* = 3). Unpaired two-tailed Student’s t-test. **(C)** Predicted FOXP2-binding motif. Schematic representation of the predicted FOXP2-binding sequence including the rs4263114 SNP within Enh-*MYCN*. **(D and E)** FOXP2 preferentially binds the non-risk allele at rs4263114. (D) Anti-FOXP2 chromatin immunoprecipitation (ChIP)-qPCR assay validating FOXP2 enrichment at the Enh-*MYCN* locus in heterozygous (T/G) cNCCs. Normal IgG served as the negative control. Mean ± SD (*n* = 3). Two-way ANOVA (Sidak’s post-hoc test). **(F)** The risk allele abolishes FOXP2-mediated transactivation of Enh-*MYCN*. Relative luciferase activity of Enh-*MYCN* reporter constructs harboring either the non-risk (T) or risk (G) allele in HEPM cells co-transfected with OE- FOXP2 or OE-Mock. Mean ± SD (n = 3). One-way ANOVA (Tukey’s post-hoc test).

To experimentally validate the allele-specific binding of FOXP2, we performed anti-FOXP2 chromatin immunoprecipitation (ChIP) in lysates from heterozygous cNCCs. A threefold enrichment of the non-risk allele over the risk allele was investigated in subsequent ChIP-qPCR analysis of the IP DNA (Figure 5D), confirming that FOXP2 preferentially occupies the non-risk sequence. Given that FOXP1 overexpression also upregulated *MYCN* (Figure S6A), we conducted parallel anti-FOXP1 ChIP-qPCR assays (Figure S6C). In contrast to FOXP2, FOXP1 showed no allelic binding bias at the rs4263114 locus, supporting the specificity of the FOXP2-enhancer interaction.

Furthermore, dual-luciferase assays in HEPM cells demonstrated that upregulating FOXP2 robustly increased the activity of Enh-*MYCN* carrying the non-risk sequence, however, the presence of the rs4263114 risk variant entirely neutralized this transactivation effect (Figure 5E). These results indicate that the rs4263114 risk allele disrupts the recruitment of FOXP2 to Enh-*MYCN*, thereby suppressing *MYCN* expression.

### Risk allele of rs4263114 reduces *MYCN* expression by compromising FOXP2 liquid-liquid phase separation

We hypothesized that FOXP2 modulates *MYCN* transcription through LLPS and that the risk allele perturbs this process. Bioinformatic analysis using PONDR and PLAAC revealed that FOXP2 contains a prominent N-terminal IDR (aa. 101-232), providing a structural basis for phase separation (Figures 6A and 6B). Consistent with this, PSPHunter predicted a high phase-separation propensity for FOXP2, with a score of 0.6225 (Figure S7A).

**Figure 6.**
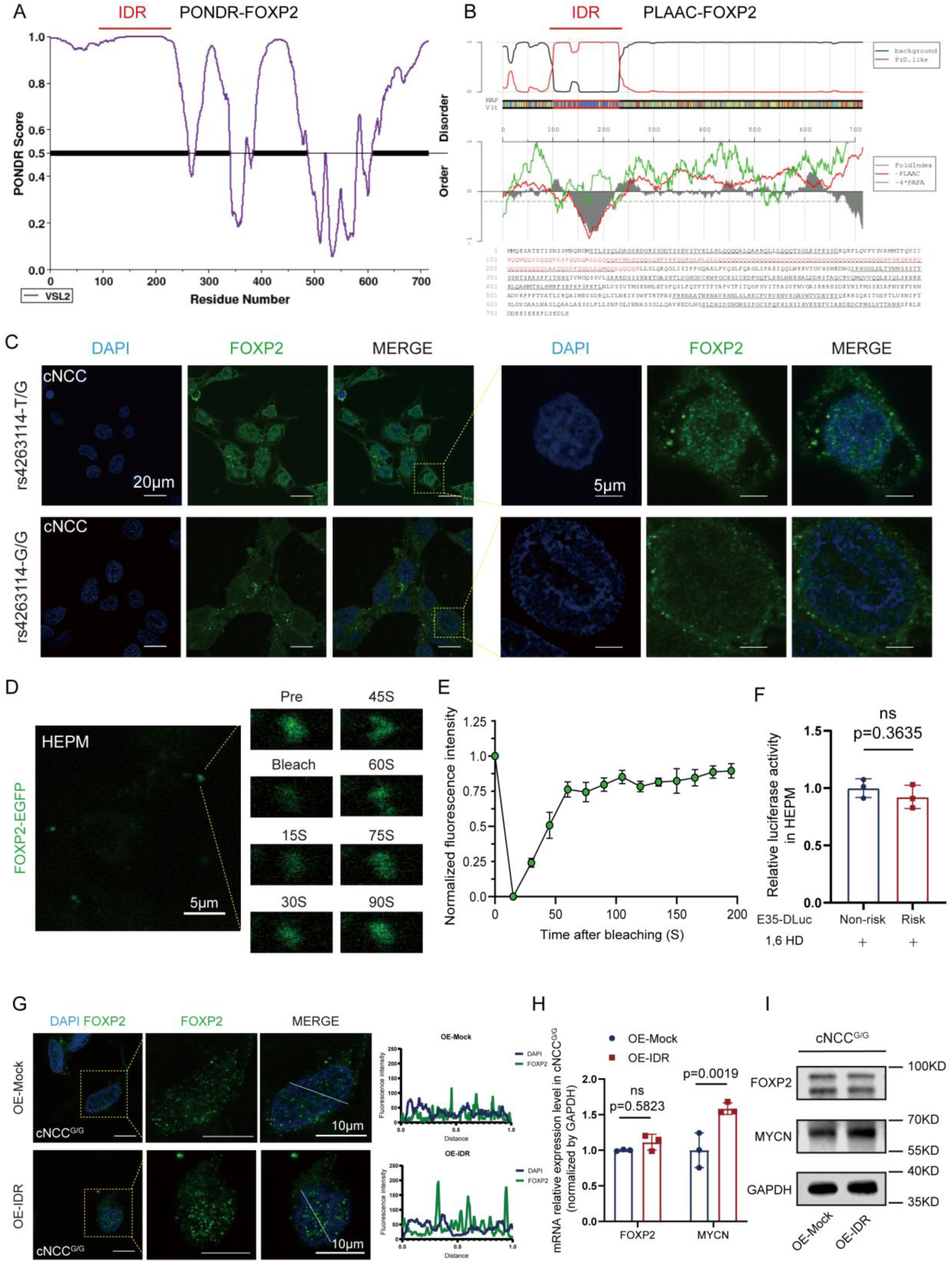
The rs4263114 risk allele downregulates *MYCN* expression by compromising FOXP2 liquid-liquid phase separation. **(A and B)** FOXP2 contains a prominent intrinsically disordered region (IDR). Bioinformatic prediction of the FOXP2 IDR sequence using (A) PONDR and (B) PLAAC, providing the structural basis for phase separation. **(C)** The risk allele disrupts the formation of FOXP2 nuclear condensates. Representative immunofluorescence images of FOXP2 in cNCCs harboring heterozygous (T/G) or homozygous risk (G/G) alleles at rs4263114. Droplet-like FOXP2 condensates are prominent in T/G cells but show a diffuse distribution in G/G cells. Scale bars, 20 μm (left) and 5 μm (right). **(D and E)** FOXP2 condensates show highly dynamic liquid-like properties. (D) Representative time-lapse fluorescence recovery after photobleaching (FRAP) images of EGFP-FOXP2 overexpressing HEPM cells. Scale bar, 5 μm. (E) Quantification of normalized fluorescence recovery intensity over time (*n* = 3 independent experiments). **(F)** LLPS is required for the allele-specific regulatory activity of Enh-*MYCN*. Dual-luciferase reporter assay comparing the activity of Enh-*MYCN* constructs harboring the non-risk (T) or risk (G) allele in HEPM cells upon treatment with 1.5% 1,6-hexanediol (1,6-HD) to chemically disrupt LLPS. Mean ± SD (*n* = 3). Unpaired two-tailed Student’s t-test. **(G-I)** Overexpression of the FOXP2-IDR domain rescues phase separation and *MYCN* expression in risk-allele cNCCs. (G) Immunofluorescence staining of FOXP2 showing restoration of nuclear condensates in homozygous risk (G/G) cNCCs after FOXP2-IDR overexpression (OE-IDR) versus a Mock-vector baseline (OE-Mock). Scale bar, 10 μm. (H) RT-qPCR and (I) western blot analyses of FOXP2 and *MYCN* mRNA levels, and their corresponding protein levels, in OE-Mock and OE-IDR G/G cNCCs. Data normalized to *GAPDH*. For (H) Mean ± SD (*n* = 3). Two-way ANOVA (Sidak’s post-hoc test).

To test this hypothesis, we performed immunofluorescence staining for FOXP2 in cNCCs carrying different rs4263114 genotypes. In heterozygous cells, FOXP2 formed distinct droplet-like nuclear condensates (Figure 6C). In contrast, in cells harboring the homozygous risk variant, FOXP2 showed a diffuse and uniform fluorescence pattern throughout the nucleus (Figure 6C). These observations suggest that the risk allele disrupts initial condensate formation of *FOXP2*.

To confirm the liquid-like properties of these condensates, expression vectors carrying either EGFP-FOXP2 or unfused EGFP were introduced into HEPM cells. In the nuclei of HEPM cells, EGFP-FOXP2 assembled into distinct droplet-like condensates, whereas EGFP alone exhibited a diffuse fluorescence pattern (Figure S7B). We then performed FRAP in HEPM cells overexpressing EGFP-FOXP2. The bleached EGFP-FOXP2 condensates showed rapid fluorescence recovery within 100 s, indicating highly dynamic liquid-like behavior (Figures 6D and 6E; Supplemental Video). The liquid-like nature of these FOXP2 condensates was additionally confirmed by real-time cellular monitoring, which revealed their capacity to undergo unprompted coalescence. (Figure S7C). To corroborate this phase-separation behavior *in vivo*, the EGFP-FOXP2 mRNA was microinjected into single-cell zebrafish zygotes, which further validating the capacity of FOXP2 to undergo spontaneous phase separation. (Figure S7D).

To determine whether phase separation is required for transcriptional activation, we monitored reporter activity in HEPM cells using dual-luciferase assays following the administration of 1,6-hexanediol (1,6-HD). This chemical probe specifically abolishes LLPS by impairing essential hydrophobic contacts.^51^ Strikingly, 1,6-HD treatment abolished the higher activity of the non-risk sequence (T) relative to the risk sequence (G) (Figure 6F). This result indicates that LLPS is required for the allele-specific regulatory activity of Enh-*MYCN*. Finally, we performed rescue experiments. Because the IDR (aa. 101-232) is crucial for LLPS (Figures 6A and 6B), we overexpressed the FOXP2-IDR domain in homozygous risk (G/G) cNCCs. This overexpression increased the formation of droplet-like condensates (Figure 6G). In the homozygous risk (G/G) group, FOXP2-IDR overexpression also partially restored *MYCN* expression (Figures 6H and 6I). These findings demonstrate that the rs4263114 risk allele impairs *MYCN* expression by disrupting FOXP2 LLPS, providing a new epigenetic mechanism underlying NSCLP susceptibility.

## Discussion

As the top susceptibility locus for NSCLP and the third susceptibility locus for NSOFC, functionally annotating 2p24.2 remains essential for understanding disease origins. Therefore, our research sought to identify specific causal variants alongside local regulatory elements, ultimately revealing molecular mechanisms driving such widespread congenital defects. Together with previously characterized risk loci,^13,31,52,53^ these findings provide a basis for translating genetic discoveries into more precise clinical genetic counseling.

Building on the regulatory framework provided by FaceBase^49^ and the prior identification of the *DDX1* enhancer (e2p24.2),^33^ we conducted systematic genetic screening in a Chinese cohort comprising 2,437 cases and 2,391 controls. This approach prioritized rs4263114 as a functional variant located within a novel enhancer, Enh-*MYCN*. Population frequencies from gnomAD v4.1.0 show that the risk allele is substantially enriched in East Asians (AF = 0.7225) relative to non-Finnish Europeans (AF = 0.2878). This disparity raises the possibility that different causal variants within this region contribute to NSCLP susceptibility in European populations, highlighting the need to further explore the genetic heterogeneity of NSCLP across ancestries.

Previous studies have established *MYCN* as a key regulator of craniofacial development.^54,55^ By integrating cNCCs, zebrafish models, and human mandibular organoids, we systematically dissected the regulatory mechanism of *MYCN* in both cellular and complex three-dimensional tissue contexts. We show that the rs4263114 risk allele downregulates *MYCN* expression and directly impairs the osteogenic differentiation capacity of cNCCs. Overall, our data confirm that *MYCN* expression levels are essential for normal craniofacial development.

Recent advances in chromatin biology have shown that LLPS facilitates DNA-protein interactions to drive enhancer function.^56–59^ Active enhancers recruit TFs and coactivators to form dynamic condensates known as transcription hubs.^57,58,60^ TFs containing IDRs are major drivers of this phase-separation process,^61^ ensuring efficient and spatiotemporal gene regulation.^51,61,62^ As a Forkhead family member, FOXP2 is a key regulator of organogenesis and craniofacial development.^50,63,64^ Emerging evidence suggests that Forkhead proteins, such as FOXP1 (*FOXP1* [MIM: 605515]) and FOXP2, have phase-separating properties.^65,66^ Our data indicate that disruption of FOXP2 phase separation in cells carrying the homozygous risk allele is not incidental. Rather, the rs4263114 variant reduces FOXP2-binding affinity, thereby impairing the local accumulation of FOXP2 condensates and weakening long-range regulation of *MYCN*. This mechanism parallels recent findings in complex disease genetics,^56,67,68^ such as RUNX2-mediated regulation of *XCR1* (*XCR1* [MIM: 600552]) in osteoporosis^56^ and ARID3B-mediated regulation in NSCLP.^67^ Thus, FOXP2 is identified here as a TF that modulates non-coding disease-risk variants through an LLPS-dependent mechanism.

We acknowledge several limitations of this study. First, because genetic heterogeneity across populations is common in the etiology of OFCs,^69^ findings derived exclusively from the Han Chinese population may not fully capture the risk landscape in other ancestries. The potential causal variants in other populations remain to be identified. Second, for another linked variant, rs4240234 within Enh-*MYCN*, technical constraints prevented systematic dissection of its independent function. Notably, a recent comprehensive functional analysis of complex traits showed that adjacent variants within the same CRE frequently exhibit non-additive regulatory epistasis, jointly dampening or amplifying transcriptional output.^70^ Based on this concept and the established multi-hit model,^71^ we speculate that rs4240234 acts as a co-contributor that modulates enhancer activity together with the dominant driver rs4263114. The precise epistatic and synergistic effects between these closely linked variants require further investigation. Third, although the current model supports a mechanism in which FOXP2-mediated phase separation drives enhancer-promoter looping, transcriptional regulation is inherently multimodal. Emerging evidence suggests that other genomic features, such as enhancer RNAs (eRNAs),^72,73^ *Alu* elements,^74^ or short tandem repeats (STRs),^75^ also participate in enhancer-promoter regulation. Determining whether these mechanisms contribute to the long-range regulation of *MYCN* by distal enhancers at 2p24.2 will be necessary for a more complete understanding of NSCLP etiology.

In summary, we identified a functional non-coding variant, rs4263114, within Enh-*MYCN* at 2p24.2. Mechanistically, the risk allele downregulates *MYCN* expression and impairs cNCC differentiation by disrupting *FOXP2* phase separation, thereby conferring susceptibility to NSCLP. This work not only functionally annotates a leading GWAS locus but also, together with risk variants characterized at other loci, advances the field toward more precise genetic counseling. It also provides a conceptual framework for future risk assessment studies.

## Data availability

The datasets and experimental resources reported here are obtainable from the corresponding author upon reasonable inquiry. Additionally, the targeted sequence information is hosted by the Genome Sequence Archive (GSA) (http://gsa.big.ac.cn) and can be accessed via BioProject PRJCA059597.

## Acknowledgments

We are profoundly grateful to Prof. Robert A. Cornell (University of Washington) for his critical reading and invaluable intellectual insights of the manuscript.

This work was supported by the Joint Funds of the National Natural Science Foundation of China (Grant No. U22A20313 to M.H.), the National Natural Science Foundation of China (Grant No.82170944, 81970923, and 82370966 to Z.B.; Grant No.82571053, 81970904 to M.H.; Grant No.82501089 to Z.H.), the Research Project of School and Hospital of Stomatology Wuhan University (No. ZW202401 to M.H.), the Key R&D projects of Hubei Provincial Science and Technology Plan (No. 2023BCB134 to M.H.), the Hubei Provincial Science and Technology Innovation Base (Platform) Project (Grant No. 2024CSA065 to Z.B.).

## Author contributions

Z.W., R.Y., Z.B., and M.H. conceptualized the study. Z.W. performed all experiments and analyses if not otherwise noted. Z.W wrote the manuscript. Z.Y. provided assistance in cell culture, cell behavioral analyses and RT-qPCR. R.Y. offered assistance in zebrafish microinjection. Z.H. helped develop experiments in dual luciferase assay and cell behavioral analyses. Y.L. offered support in plasmid construction. L.S. provided assistance in sequencing and data analyses. Z.B. and M.H. provided the lab space, cohort data and samples, and supervised the study. Z.H., Z.B. and M.H. acquired funding. M.H. edited the manuscript and helped develop experiments and analyses.

## Web resources

NHGRI-EBI GWAS Catalog, www.ebi.ac.uk/gwas

OMIM, http://www.omim.org/

Adobe Illustrator, https://www.adobe.com/products/illustrator

PLINK version 1.07 software, http://pngu.mgh.harvard.edu/purcell/plink/

fastx tool, http://hannonlab.cshl.edu/fastx_toolkit/index.html

FaceBase, https://www.facebase.org/

CRISPRscan algorithm, http://www.crisprscan.org/

3D Genome Browser 2.0, https://3dgenome.fsm.northwestern.edu/

Fiji (ImageJ), https://fiji.sc/

JASPAR, http://jaspar.genereg.net/

PONDR, https://www.pondr.com/

PLAAC, http://plaac.wi.mit.edu/

PSPHunter, http://psphunter.stemcellding.org/

genomAD, https://gnomad.broadinstitute.org/

