## Supplemental Information for "A non-coding variant at 2p24.2 confers susceptibility to non-syndromic cleft lip and palate through LLPS-dependent regulation of *MYCN*"

### Supplemental Figures

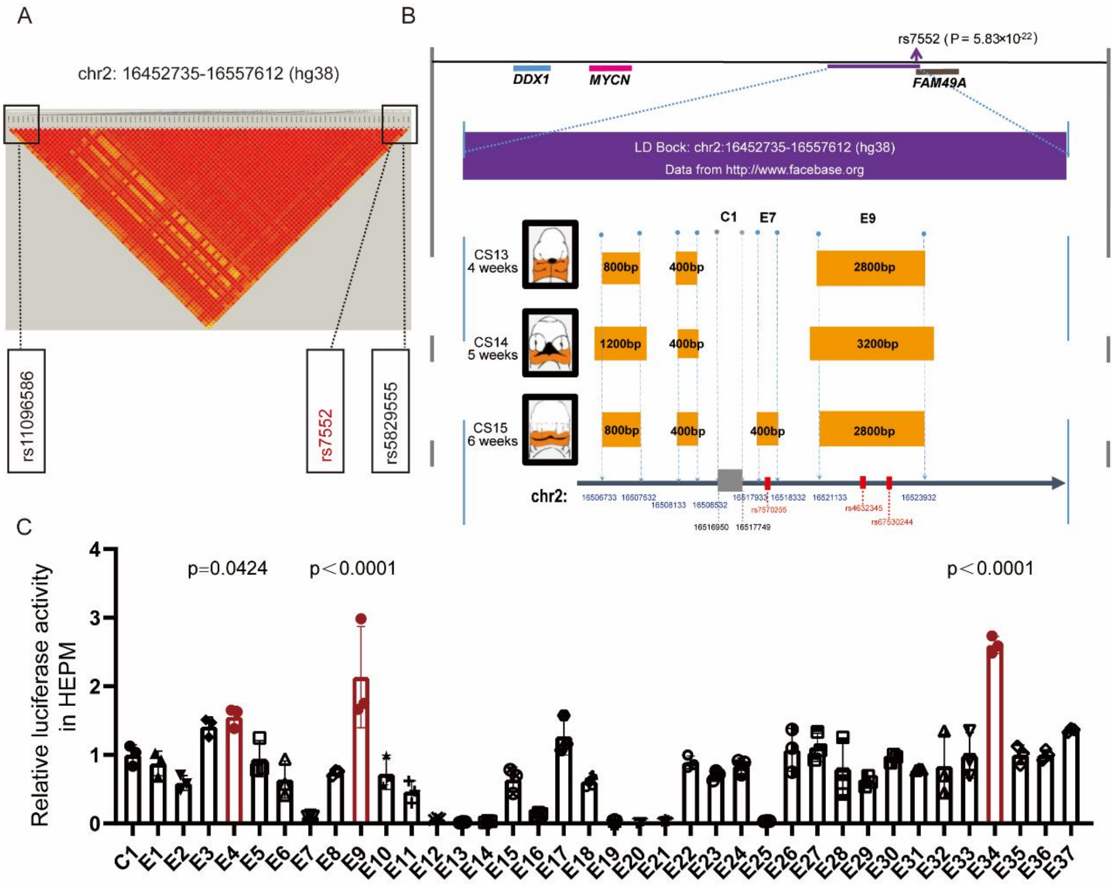

**Figure S1. Linkage disequilibrium mapping, epigenomic annotation, and supplementary functional characterization of candidate elements.**

**(A)** Delineation of the linkage disequilibrium (LD) block. The LD block surrounding the tag SNP rs7552 at the 2p24.2 locus was calculated and visualized using PLINK.

**(B)** Epigenomic annotation identifies candidate enhancers and a negative control region. Genomic tracks display three candidate SNPs (red dots) overlapping active enhancers (orange boxes) annotated in human embryonic craniofacial tissues at Carnegie stages 13-15 (CS13-CS15) from the FaceBase database. The defined LD block is indicated by a purple line. The negative control fragment, C1 (gray box), is located in a genomic region lacking active enhancer signatures. Protein-coding genes within the topologically associating domain (TAD), including *DDX1* (blue line), *MYCN* (pink line), and *FAM49A* (brown line), are shown.

**(C)** Comprehensive *in vitro* screening of candidate enhancers. Among all cloned elements, relative luciferase activities were evaluated by dual-luciferase reporter assays in HEPM cells. Mean ± SD ($n$ = 3). Two-way ANOVA (Dunnett's post hoc test).

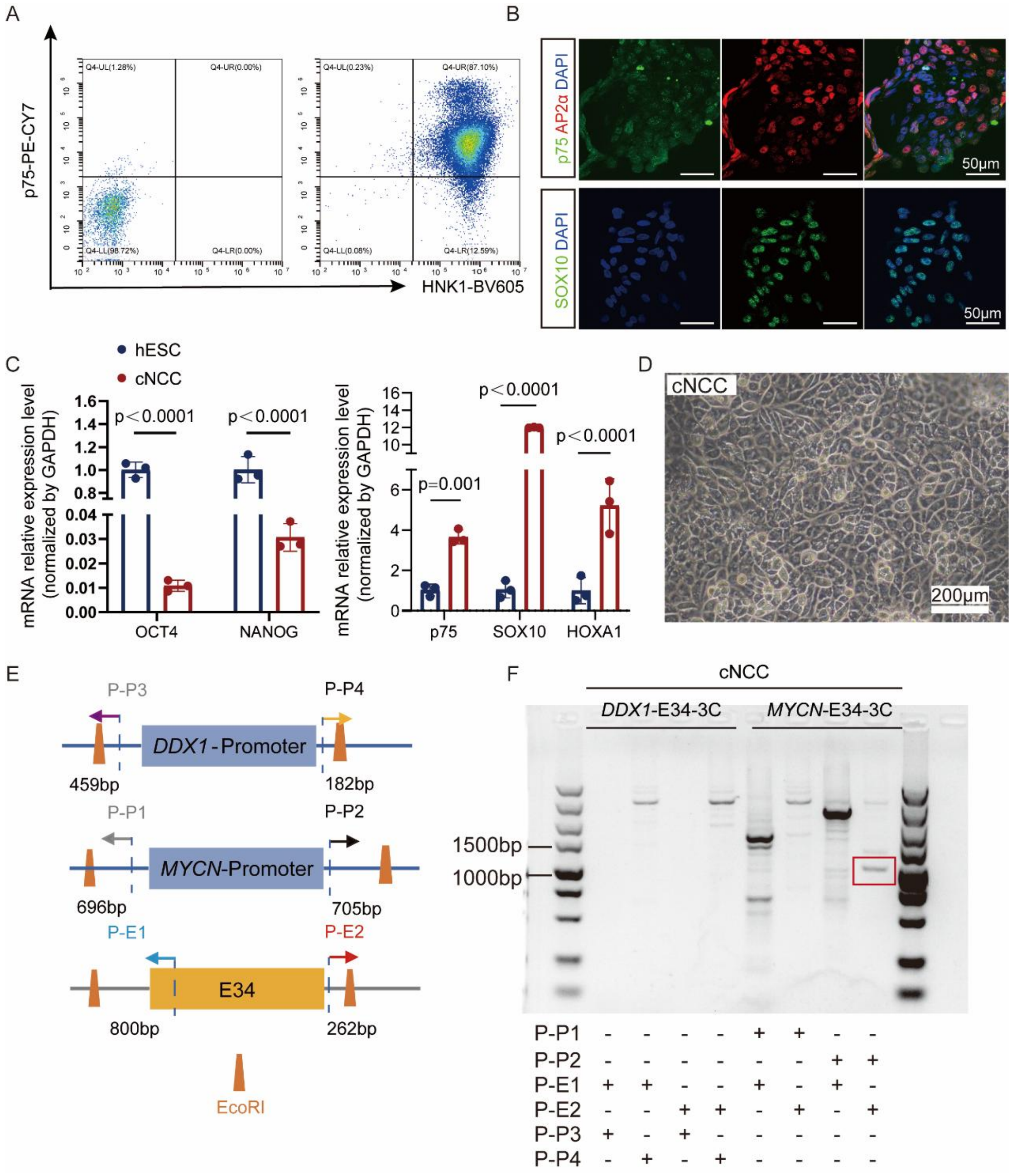

**Figure S2. Enh-*MYCN* (E34) specifically interacts with the *MYCN* promoter in thoroughly characterized hESC-derived cNCCs.**

**(A-D)** Characterization and validation of hESC-derived cNCCs. **(A)** Flow-cytometric analysis demonstrating that for the cNCC-specific surface markers p75 and HNK-1, double-positive differentiated cells reach nearly 90%. **(B)** Representative immunofluorescence images confirming robust protein expression of the cNCC markers *NGFR (p75)*, *SOX10*, and *AP2α*.

**(C)** RT-qPCR analysis showing significant upregulation of neural crest marker genes (*p75*, *SOX10*, and *HOXA1*) and concomitant downregulation of pluripotency markers (*OCT4* and *NANOG*) during differentiation. Data normalized to *GAPDH*. Mean ± SD ($n$ = 3). Two-way ANOVA (Sidak's post hoc test). (D) Representative bright-field morphology of differentiated cNCCs. Scale bar, 200 µm.

**(E and F)** Enh-*MYCN* establishes a specific long-range interaction with the *MYCN* promoter, but not the DDX1 promoter. (E) Schematic of the chromatin conformation capture (3C) assay design. EcoRI restriction sites flanking E34, the *MYCN* promoter, and the *DDX1* promoter are indicated by orange trapezoids. Primer positions relative to the EcoRI sites are denoted by colored arrows (P-P1, 696 bp; P-P2, 705 bp; P-P3, 459 bp; P-P4, 182 bp; P-E1, 800 bp; and P-E2, 262 bp upstream of the respective restriction sites). (F) Agarose gel electrophoresis of 3C-PCR products. A specific 967-bp amplification product (red box) was detected using the P-P2 and P-E2 primer pair, linking E34 and the *MYCN* promoter, whereas no expected products were observed using the *DDX1* promoter-specific primer pairs.

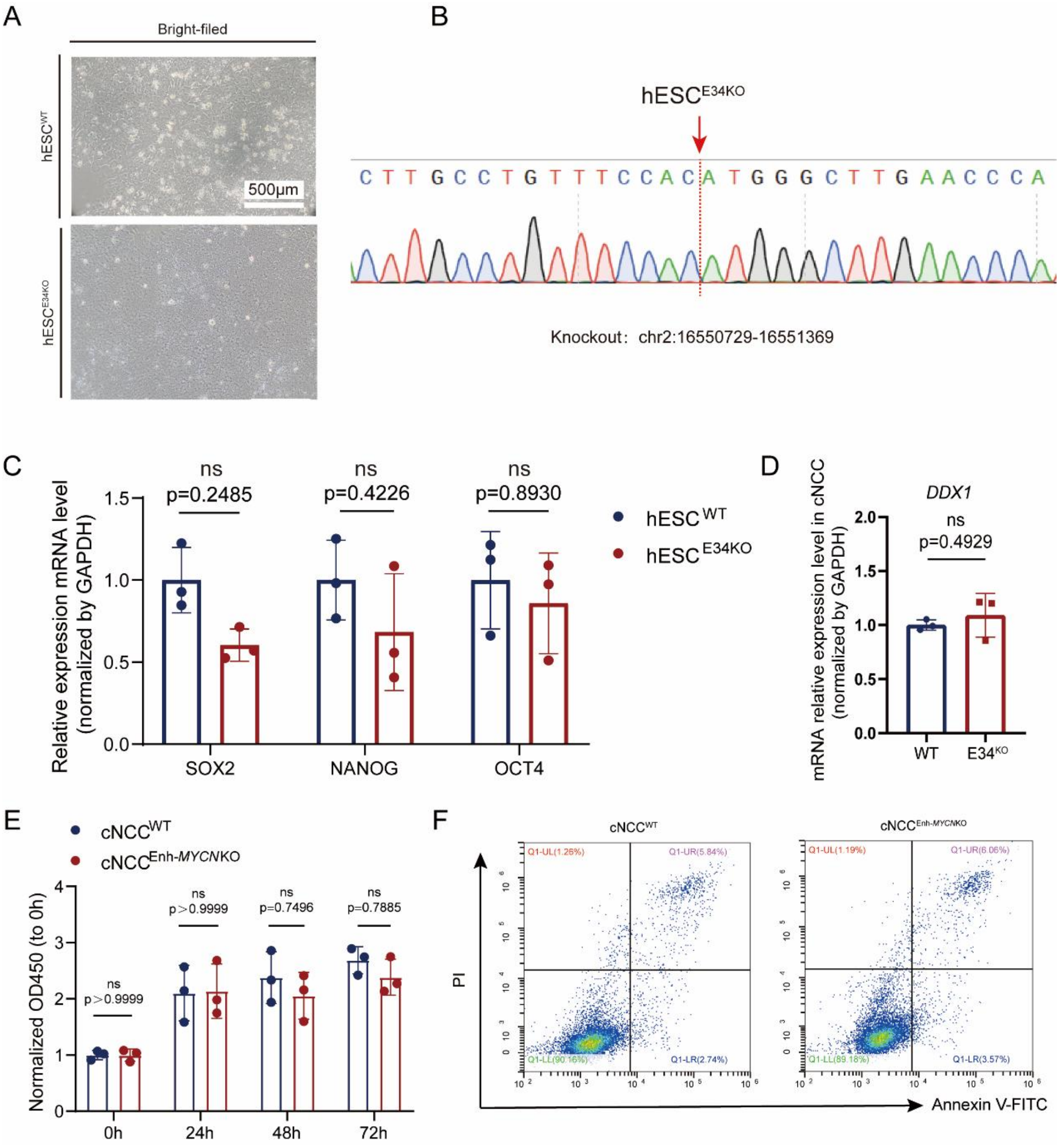

**Figure S3. Validation of the isogenic Enh-*MYCN* (E34) knockout hESC line and assessment of basal cellular functions.**

**(A-C)** Generation and characterization of the isogenic Enh-*MYCN* knockout hESC line. **(A)** Representative bright-field images showing the typical morphology of wild-type (hESC$^{WT}$) and knockout (hESC$^{Enh\text{-}MYCN\ KO}$) colonies. Scale bar, 500 μm. **(B)** Sanger sequencing chromatogram confirming the precise deletion of the target genomic region (chr2:16,550,729-16,551,369) in the hESC$^{Enh\text{-}MYCN\ KO}$ line. **(C)** RT-qPCR analysis demonstrating that expression of the core pluripotency markers *SOX2*, *NANOG*, and *OCT4* remained unchanged after enhancer deletion. Data normalized to *GAPDH*. Mean ± SD (*n* = 3). Two-way ANOVA (Sidak's post hoc test).

**(D)** Deletion of Enh-*MYCN* does not alter *DDX1* expression. RT-qPCR analysis of *DDX1* mRNA levels in $cNCC^{WT}$ and $cNCC^{Enh\text{-}MYCN\ KO}$ cells. *GAPDH* was used for normalization. Mean ± SD (*n* = 3). Unpaired two-tailed Student's t-test.

**(E and F)** Enh-*MYCN* deficiency does not affect cNCC proliferation or apoptosis. (E) Proliferation capacities of $cNCC^{WT}$ and $cNCC^{Enh\text{-}MYCN\ KO}$ cells remained statistically indistinguishable during the log phase. Mean ± SD (*n* = 3). Two-way ANOVA (Sidak's post hoc test). (F) Flow-cytometric analysis using Annexin V/PI double staining demonstrating that Enh-*MYCN* knockout does not induce apoptosis in cNCCs.

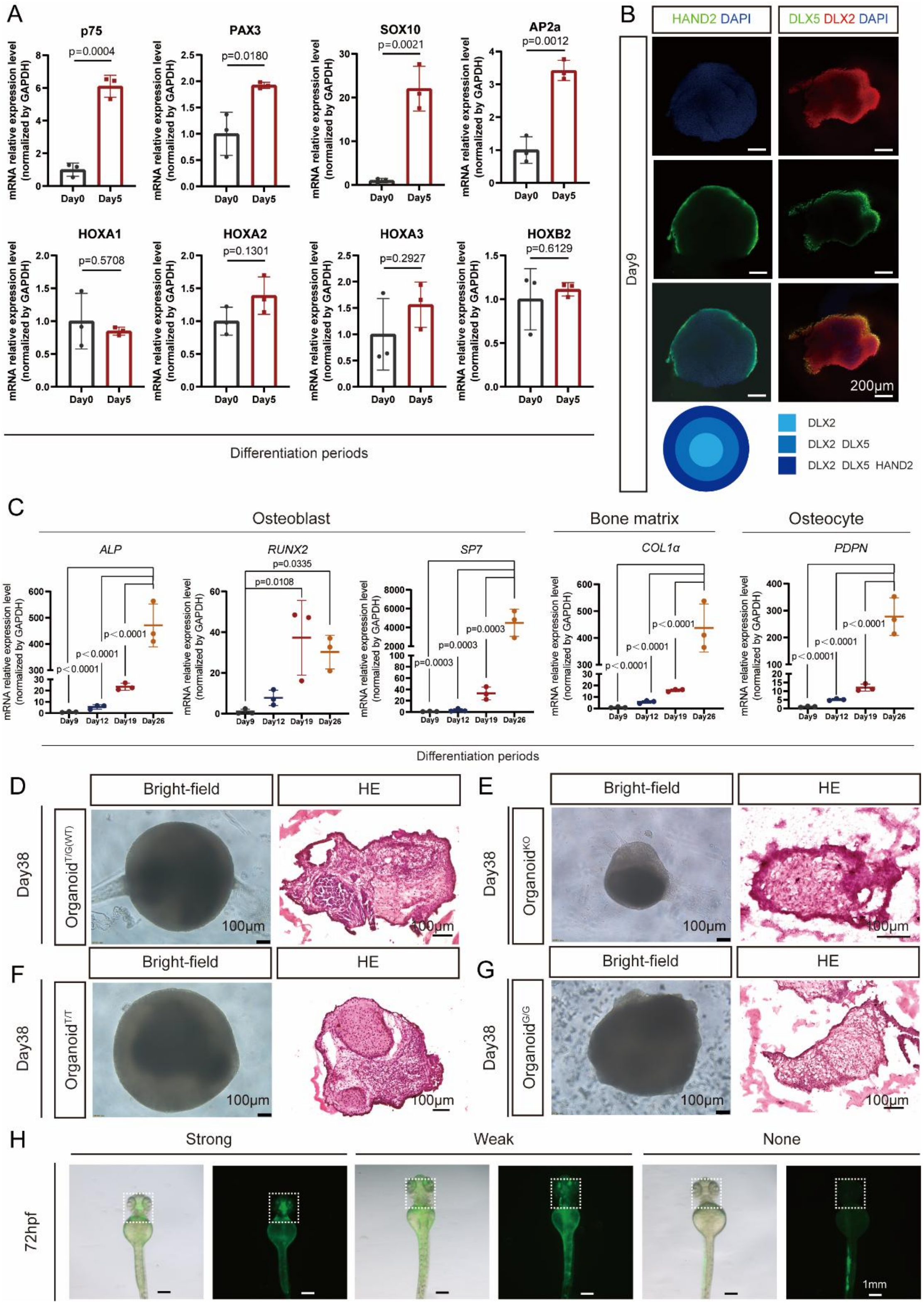

62

63 **Figure S4. Stepwise characterization of jawbone-like organoids and histological**

64 **evaluation of genotype-specific morphological defects.**

65 **(A-C)** Stepwise validation of human mandibular organoid differentiation. (A) RT-qPCR

analysis at day 5 showing successful induction of HOX-negative cNCCs. The cells exhibited robust expression of neural crest markers *NGFR* (*p75*), *PAX3*, *SOX10*, and *TFAP2A* (*AP2α*), with no expression of HOX family genes (*HOXA1*, *HOXA2*, *HOXA3*, and *HOXB2*). Data normalized to *GAPDH*. Mean ± SD ($n$ = 3). Unpaired two-tailed Student's t-test. (B) Representative immunofluorescence images at day 9 showing region-specific expression of mandibular prominence ectomesenchyme (mdEM) markers *HAND2*, *DLX2*, and *DLX5*. (C) RT-qPCR analysis tracking the temporal expression dynamics of osteoblast (*ALP*, *RUNX2*, and *SP7*), bone matrix (*COL1A1*), and mature osteocyte (*PDPN*) markers across days 5, 9, 19, and 38 of differentiation. Data normalized to *GAPDH*. Mean ± SD ($n$ = 3). One-way ANOVA (Tukey's post hoc test).

**(D-G)** Enh-*MYCN* knockout and the rs4263114 risk allele impair jawbone-like organoid morphogenesis and matrix deposition. Representative bright-field images (left) and hematoxylin and eosin (H&E) staining (right) of day 38 (d38) mandibular organoids derived from (D) hESC$^{T/G\ (WT)}$, (E) hESC$^{Enh\text{-}\mathit{MYCN\ KO}}$, (F) hESC$^{T/T}$, and (G) hESC$^{G/G}$ lines. Morphological and histological evaluation showed that Enh-*MYCN* KO organoids were globally smaller than WT organoids. Furthermore, organoids carrying the homozygous risk allele (G/G) exhibited a notably looser tissue architecture and reduced bone matrix deposition compared with their T/T and T/G counterparts. Scale bars, 100 μm.

**(H)** Grading system for *in vivo* zebrafish reporter assays. Representative fluorescence images illustrating the classification criteria (strong, weak, or none) for GFP expression in the pharyngeal arches of zebrafish embryos at 72 hpf. Scale bar, 1 mm.

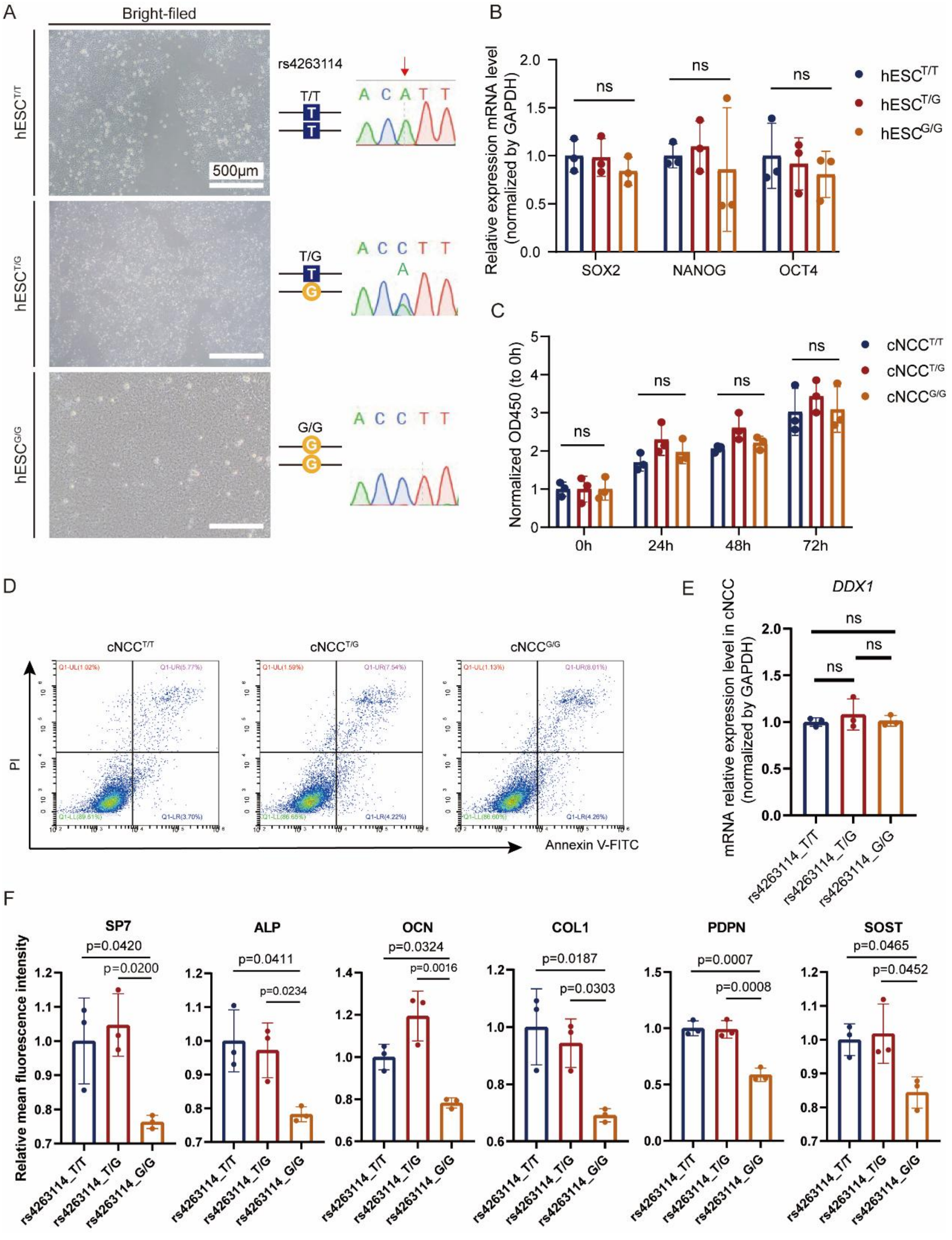

**Figure S5. Generation, characterization, and phenotypic quantification of the isogenic rs4263114 mutant hESC lines.**

**(A and B)** Generation and pluripotency validation of the isogenic rs4263114 mutant hESC lines.

(A) Representative bright-field morphology and Sanger sequencing chromatograms confirming establishment of the CRISPR/Cas9-edited $hESC^{T/T}$, $hESC^{T/G}$, and $hESC^{G/G}$ lines. Scale bar, 500 µm. (B) RT-qPCR analysis confirming that expression of the core pluripotency markers *SOX2*, *NANOG*, and *OCT4* remained comparable across the three isogenic cell lines. Data normalized to *GAPDH*. Mean ± SD ($n$ = 3). Two-way ANOVA (Tukey's post hoc test).

**(C and D)** The rs4263114 risk allele does not alter cNCC proliferation or apoptosis. (C) Proliferation capacities among $cNCC^{T/T}$, $cNCC^{T/G}$, and $cNCC^{G/G}$ remained statistically indistinguishable during their active growth phase. Mean ± SD ($n$ = 3). Two-way ANOVA (Tukey's post hoc test). (D) Flow-cytometric analysis using Annexin V/PI double staining demonstrating that the rs4263114 mutation does not induce apoptosis in cNCCs.

**(E)** The rs4263114 variant does not affect *DDX1* expression. RT-qPCR analysis of *DDX1* mRNA levels in $cNCC^{T/T}$, $cNCC^{T/G}$, and $cNCC^{G/G}$ cells. Data normalized to *GAPDH*. Mean ± SD ($n$ = 3). One-way ANOVA (Tukey's post hoc test).

**(F)** The homozygous risk allele (G/G) significantly impairs expression of osteogenic markers in mandibular organoids. Quantitative analysis of immunofluorescence staining for osteoblast and mature osteocyte markers in d38 jawbone-like organoids derived from the three isogenic lines. Mean fluorescence intensity was quantified using ImageJ, revealing a significant reduction in the G/G group compared with the T/T and T/G groups. Mean ± SD ($n$ = 3). One-way ANOVA (Tukey's post hoc test).

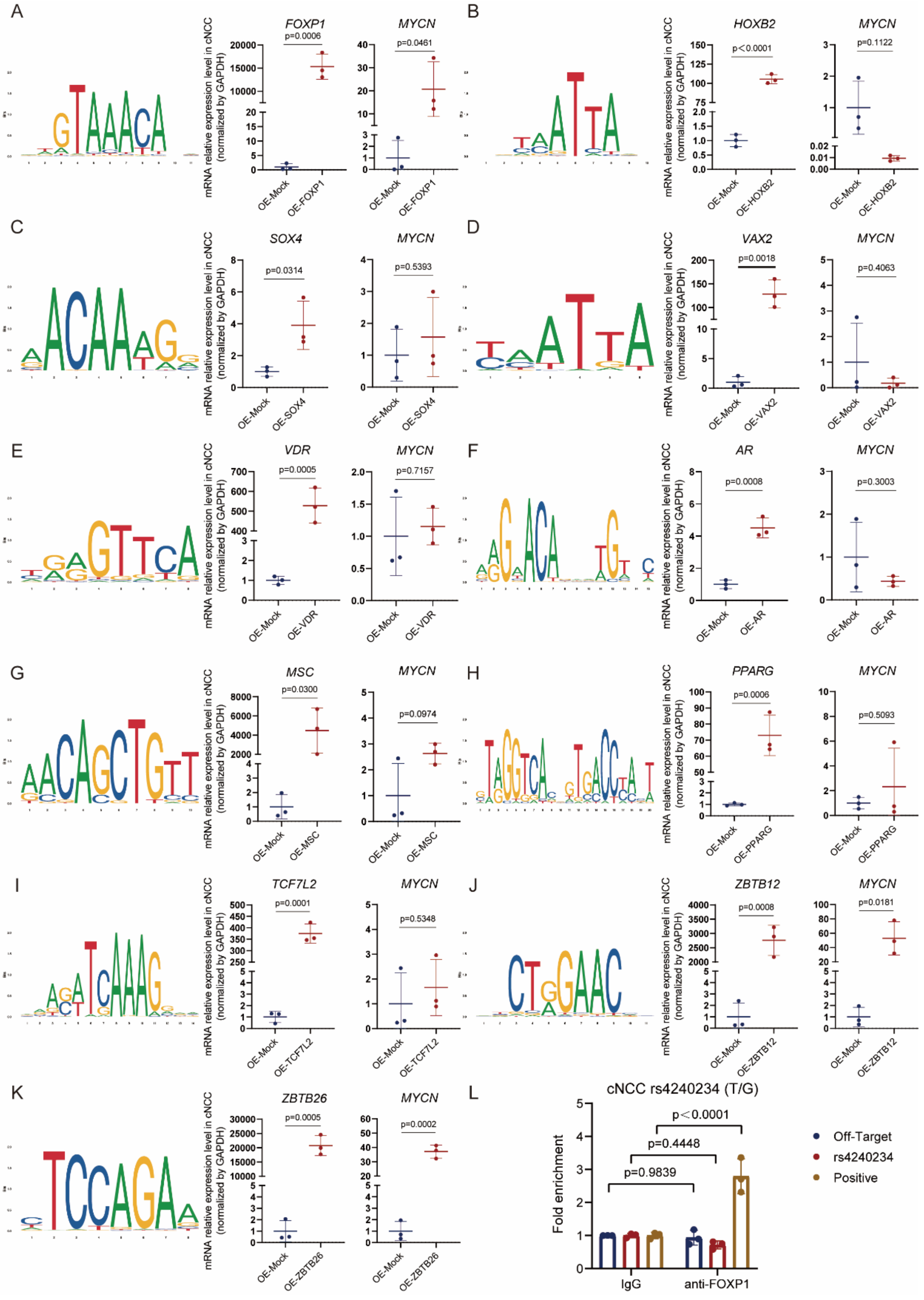

**Figure S6. Systematic functional screening and validation of predicted candidate transcription factors for the rs4263114 and rs4240234 variants.**

**(A-E)** Functional screening of candidate transcription factors associated with rs4263114.

Schematic motifs of candidate transcription factors (TFs) predicted by JASPAR to be affected by the rs4263114 variant, with their functional validation. RT-qPCR analyses of the respective TF and *MYCN* mRNA levels after overexpression of (A) *FOXP1*, (B) *HOXB2*, (C) *SOX4*, (D) *VAX2*, and (E) *VDR* in wild-type cNCCs. Among these candidates, only FOXP1 overexpression significantly upregulated *MYCN* expression. Data normalized to *GAPDH*. Mean ± SD ($n$ = 3). Unpaired two-tailed Student's t-test.

**(F-K)** Candidate TFs for rs4240234 exhibit functional profiles inconsistent with the risk enhancer defect. Schematic motifs and functional validation of candidate TFs predicted to bind the rs4240234 variant. RT-qPCR analyses of TF and *MYCN* mRNA levels after overexpression of (F) *AR*, (G) *MSC*, (H) *PPARG*, (I) *TCF7L2*, (J) *ZBTB12*, and (K) *ZBTB26* in wild-type cNCCs. Although overexpression of *ZBTB12* and *ZBTB26* promoted *MYCN* expression, this activating effect contradicts their JASPAR-predicted preferential binding to the risk allele (G), which is known to suppress rather than enhance enhancer function. Data normalized to *GAPDH*. Mean ± SD ($n$ = 3). Unpaired two-tailed Student's t-test.

**(L)** FOXP1 lacks allele-specific enrichment at rs4263114. Anti-FOXP1 chromatin immunoprecipitation (ChIP)-qPCR assay in heterozygous (T/G) cNCCs showing no significant allele-specific binding preference of FOXP1 for the rs4263114 variant. Normal IgG served as the negative control. Mean ± SD ($n$ = 3). Two-way ANOVA (Sidak's post hoc test).

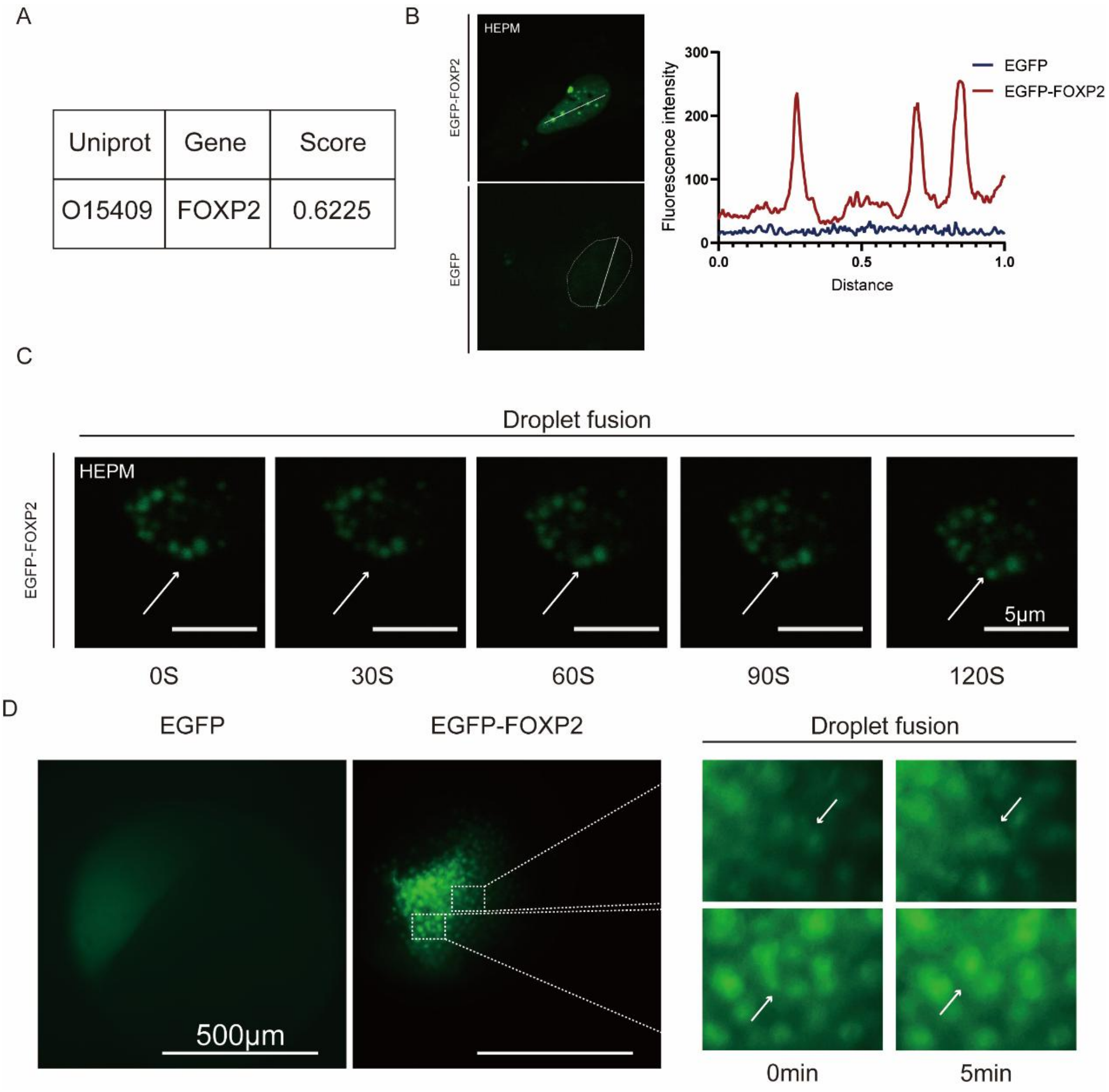

**Figure S7. Predictive and visual validation of FOXP2 liquid-liquid phase separation *in vitro* and *in vivo*.**

**(A)** Bioinformatic prediction of FOXP2 phase separation. PSPHunter analysis predicted a high propensity of the FOXP2 protein to undergo liquid-liquid phase separation (LLPS).

**(B and C)** FOXP2 forms dynamic liquid-like condensates *in vitro*. (B) Representative confocal images of HEPM cells overexpressing either EGFP (control) or EGFP-FOXP2. Scale bar, 5 μm. The right panel shows the corresponding quantification of fluorescence intensity along the indicated cross-sectional line (white), highlighting the discrete, highly concentrated distribution of FOXP2 droplets. (C) Time-lapse live-cell imaging capturing spontaneous fusion of two adjacent EGFP-FOXP2 droplets in HEPM cells over 120 s, a hallmark of liquid-like behavior. Scale bar, 5 μm.

**(D)** FOXP2 undergoes phase separation *in vivo*. Live-cell confocal imaging showing that EGFP-FOXP2 forms self-assembled granular condensates (green) in developing zebrafish embryos at 2 hpf. Scale bar, 500 μm.

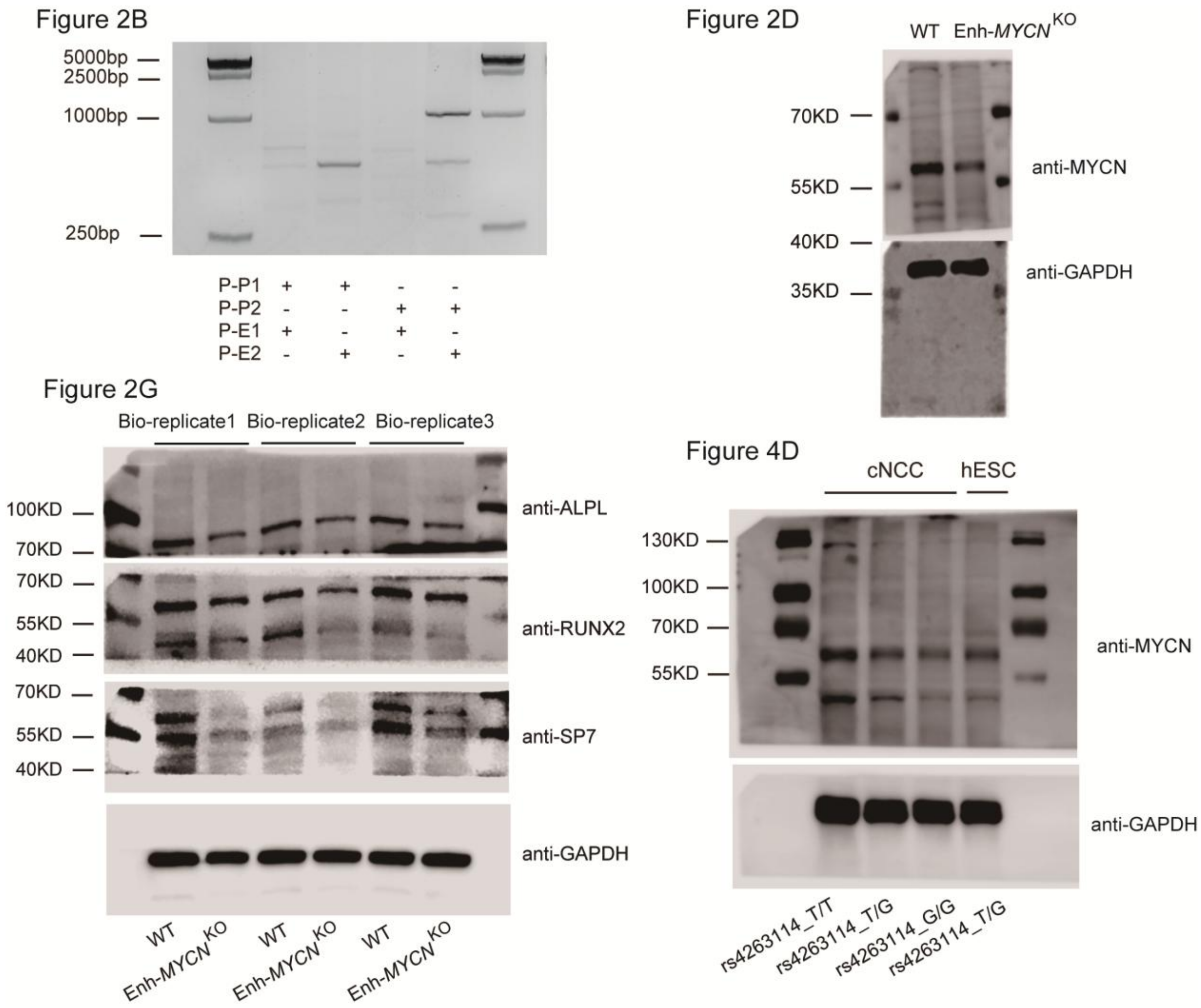

**Figure S8. The uncropped blots or gel in this study.**

**Figure S9. The uncropped blots in this study.**

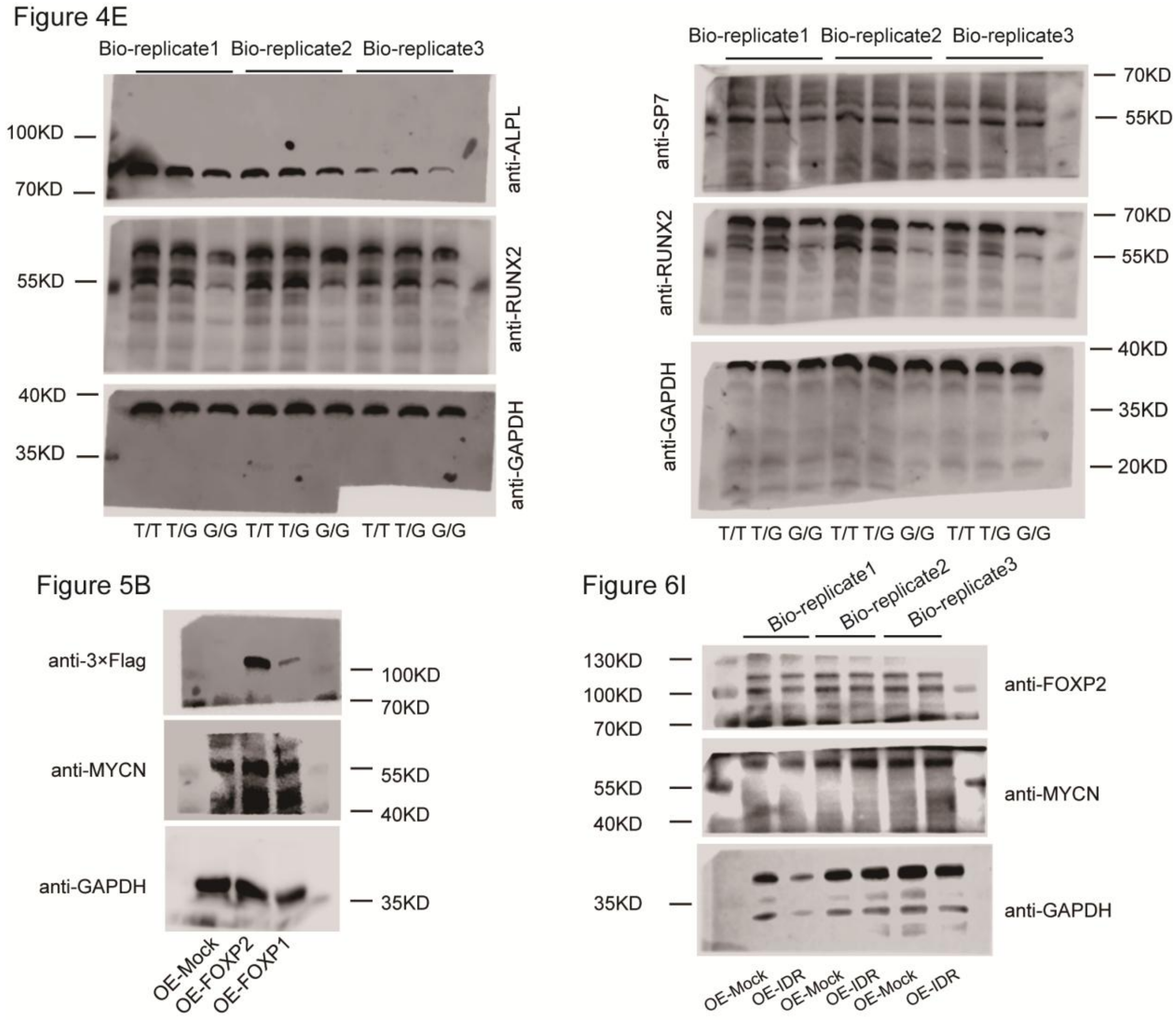

### Supplemental Tables

**Table S1: Samples summary in the targeted sequencing and replication studies**

| Stage | Cohort | Ethnicity | Phenotype | Cases | | Controls | |
|---|---|---|---|---|---|---|---|
| | | | | Sample size | Male/Female (%) | Sample size | Male/Female (%) |
| Discovery | cohort 1 | Chinese | NSCLP[a] | 1414 | 72.7/27.3 | 1371 | 57.8/42.2 |
| Replication | cohort 2 | Chinese | NSCLP | 1025 | 72.5/27.5 | 1022 | 50.1/49.9 |

NSCLP: non-syndromic cleft lip with palate

**Table S2: Association results for SNPs from the 2p24.2 risk loci at each stage**

| SNP | BP (hg38) | risk allele/non-risk allele | Discovery (above) and Replication (below) [a] | | | | Meta | | |
|---|---|---|---|---|---|---|---|---|---|
| | | | F_A[b] | F_U[c] | *P* | OR[d] | *P* | OR | *P*HET[e] |
| rs11096586 | 2:16476564 | T/C | 0.81 | 0.76 | 4.57E-04 | 1.37 | 2.09E-05 | 1.23 | 0.8063 |
| | | | 0.79 | 0.78 | 1.85E-01 | 1.11 | | | |
| rs4632344 | 2:16506696 | T/C | 0.77 | 0.71 | 3.47E-05 | 1.38 | 2.93E-07 | 1.27 | 0.7622 |
| | | | 0.74 | 0.72 | 4.18E-02 | 1.16 | | | |
| rs4490186 | 2:16506698 | C/A | 0.77 | 0.71 | 3.47E-05 | 1.38 | 2.93E-07 | 1.27 | 0.7622 |
| | | | 0.74 | 0.72 | 4.18E-02 | 1.16 | | | |
| rs34317246 | 2:16509871 | G/A | 0.78 | 0.71 | 2.06E-06 | 1.43 | 3.65E-09 | 1.32 | 0.7409 |
| | | | 0.76 | 0.72 | 8.85E-03 | 1.21 | | | |
| rs11096594 | 2:16510703 | C/A | 0.78 | 0.71 | 3.55E-06 | 1.42 | 5.47E-09 | 1.31 | 0.7269 |
| | | | 0.76 | 0.72 | 8.89E-03 | 1.21 | | | |
| rs4564771 | 2:16512560 | G/T | 0.78 | 0.71 | 2.20E-06 | 1.43 | 3.66E-09 | 1.32 | 0.7423 |
| | | | 0.76 | 0.72 | 8.87E-03 | 1.21 | | | |
| rs35165397 | 2:16514105 | C/T | 0.78 | 0.72 | 7.09E-06 | 1.42 | 8.25E-09 | 1.31 | 0.7103 |
| | | | 0.76 | 0.72 | 8.44E-03 | 1.21 | | | |
| rs7560047 | 2:16515385 | G/C | 0.78 | 0.71 | 2.65E-06 | 1.43 | 3.43E-09 | 1.32 | 0.7262 |
| | | | 0.76 | 0.72 | 7.59E-03 | 1.21 | | | |
| rs9807969 | 2:16515507 | A/G | 0.69 | 0.63 | 3.52E-04 | 1.32 | 4.29E-06 | 1.22 | 0.8253 |
| | | | 0.66 | 0.64 | 1.46E-01 | 1.10 | | | |
| rs11280766[f] | 2:16515777 | -/CATTTCAT | 0.78 | 0.71 | 2.81E-06 | 1.43 | - | - | - |
| | | | 0.76 | 0.72 | 7.98E-03 | 1.21 | | | |
| rs4334496 | 2:16516498 | G/A | 0.68 | 0.62 | 2.57E-04 | 1.33 | 6.10E-06 | 1.22 | 0.8609 |
| | | | 0.64 | 0.62 | 1.92E-01 | 1.09 | | | |
| rs6715080 | 2:16516793 | T/C | 0.78 | 0.71 | 2.19E-06 | 1.43 | 4.08E-09 | 1.32 | 0.7494 |
| | | | 0.76 | 0.72 | 9.79E-03 | 1.20 | | | |
| rs9807999 | 2:16516798 | G/A | 0.68 | 0.62 | 4.99E-04 | 1.31 | 7.47E-06 | 1.21 | 0.8307 |
| | | | 0.64 | 0.62 | 1.70E-01 | 1.10 | | | |
| rs7604116 | 2:16518083 | A/G | 0.69 | 0.63 | 3.77E-04 | 1.32 | 5.77E-06 | 1.22 | 0.8268 |
| | | | 0.66 | 0.06 | 1.59E-01 | 1.10 | | | |
| rs7570255 | 2:16518164 | T/G | 0.78 | 0.71 | 9.43E-07 | 1.45 | 3.14E-09 | 1.32 | 0.7800 |
| | | | 0.76 | 0.72 | 1.20E-02 | 1.20 | | | |
| rs11695429 | 2:16519120 | G/A | 0.78 | 0.71 | 1.02E-06 | 1.44 | 2.96E-09 | 1.32 | 0.7799 |
| | | | 0.76 | 0.72 | 1.15E-02 | 1.20 | | | |
| rs6714955 | 2:16519629 | A/C | 0.69 | 0.63 | 3.30E-04 | 1.32 | 6.52E-06 | 1.21 | 0.8450 |
| | | | 0.66 | 0.64 | 1.88E-01 | 1.09 | | | |
| rs6706465 | 2:16519820 | A/G | 0.69 | 0.63 | 3.57E-04 | 1.32 | 6.38E-06 | 1.21 | 0.8386 |
| | | | 0.66 | 0.64 | 1.77E-01 | 1.09 | | | |
| rs4632345 | 2:16521386 | G/A | 0.77 | 0.70 | 3.56E-06 | 1.42 | 2.87E-08 | 1.29 | 0.8111 |
| | | | 0.74 | 0.71 | 2.96E-02 | 1.17 | | | |

| | | | | | | | | | | |
|---|---|---|---|---|---|---|---|---|---|---|
| rs67530244 | 2:16521506 | G/A | | 0.77 | 0.71 | 8.90E-06 | 1.41 | 4.92E-08 | 1.29 | 0.7900 |
| | | | | 0.75 | 0.72 | 3.01E-02 | 1.17 | | | |
| rs75671089 | 2:16523264 | T/A | | 0.68 | 0.62 | 7.80E-04 | 1.30 | 2.55E-05 | 1.20 | 0.8662 |
| | | | | 0.64 | 0.62 | 2.98E-01 | 1.07 | | | |
| rs4832647 | 2:16524128 | G/A | | 0.77 | 0.70 | 1.31E-06 | 1.43 | 5.50E-09 | 1.31 | 0.8105 |
| | | | | 0.74 | 0.71 | 1.77E-02 | 1.18 | | | |
| rs6735186 | 2:16524646 | T/C | | 0.76 | 0.69 | 1.28E-06 | 1.43 | 2.63E-09 | 1.31 | 0.7961 |
| | | | | 0.74 | 0.70 | 1.32E-02 | 1.19 | | | |
| rs4493263 | 2:16525503 | T/C | | 0.77 | 0.70 | 4.55E-07 | 1.45 | 1.54E-09 | 1.32 | 0.8171 |
| | | | | 0.74 | 0.70 | 1.25E-02 | 1.19 | | | |
| rs4240230 | 2:16525599 | G/A | | 0.77 | 0.70 | 8.14E-07 | 1.44 | 2.48E-09 | 1.31 | 0.8118 |
| | | | | 0.74 | 0.70 | 1.36E-02 | 1.19 | | | |
| rs4497881 | 2:16525623 | A/C | | 0.77 | 0.70 | 6.57E-07 | 1.44 | 2.72E-09 | 1.31 | 0.8250 |
| | | | | 0.74 | 0.70 | 1.59E-02 | 1.18 | | | |
| rs12710725 | 2:16526254 | G/T | | 0.76 | 0.69 | 6.90E-07 | 1.44 | 1.52E-09 | 1.32 | 0.7944 |
| | | | | 0.74 | 0.70 | 9.85E-03 | 1.20 | | | |
| rs13406059 | 2:16526313 | C/G | | 0.76 | 0.69 | 6.90E-07 | 1.44 | 2.16E-09 | 1.31 | 0.8076 |
| | | | | 0.74 | 0.70 | 1.28E-02 | 1.19 | | | |
| rs4832465 | 2:16527280 | G/T | | 0.76 | 0.70 | 2.27E-06 | 1.42 | 4.54E-09 | 1.31 | 0.7860 |
| | | | | 0.74 | 0.70 | 1.31E-02 | 1.19 | | | |
| rs4832648 | 2:16527724 | T/C | | 0.78 | 0.71 | 1.18E-06 | 1.44 | 1.71E-09 | 1.32 | 0.7746 |
| | | | | 0.75 | 0.72 | 8.96E-03 | 1.20 | | | |
| rs12614326 | 2:16527879 | C/G | | 0.68 | 0.62 | 1.63E-03 | 1.29 | 2.40E-05 | 1.20 | 0.8296 |
| | | | | 0.64 | 0.62 | 2.27E-01 | 1.08 | | | |
| rs4832466 | 2:16528089 | T/A | | 0.68 | 0.62 | 1.22E-03 | 1.30 | 2.08E-05 | 1.20 | 0.8444 |
| | | | | 0.64 | 0.63 | 2.40E-01 | 1.08 | | | |
| rs6733081 | 2:16528853 | T/C | | 0.68 | 0.62 | 1.25E-03 | 1.30 | 1.97E-05 | 1.20 | 0.8388 |
| | | | | 0.65 | 0.63 | 2.30E-01 | 1.08 | | | |
| rs4832467 | 2:16529253 | C/G | | 0.68 | 0.63 | 1.69E-03 | 1.30 | 2.88E-05 | 1.20 | 0.8405 |
| | | | | 0.64 | 0.63 | 2.53E-01 | 1.08 | | | |
| rs4832468 | 2:16529297 | T/C | | 0.78 | 0.71 | 1.80E-06 | 1.43 | 2.06E-09 | 1.32 | 0.7575 |
| | | | | 0.75 | 0.71 | 8.08E-03 | 1.21 | | | |
| rs11897201 | 2:16530785 | T/C | | 0.76 | 0.69 | 2.11E-06 | 1.43 | 7.03E-09 | 1.30 | 0.7977 |
| | | | | 0.73 | 0.70 | 1.64E-02 | 1.18 | | | |
| rs6745357 | 2:16532127 | G/C | | 0.80 | 0.74 | 3.38E-05 | 1.40 | 1.89E-06 | 1.26 | 0.8784 |
| | | | | 0.77 | 0.75 | 1.75E-01 | 1.11 | | | |
| rs10205870 | 2:16532882 | G/A | | 0.67 | 0.61 | 1.24E-03 | 1.30 | 3.29E-05 | 1.19 | 0.8695 |
| | | | | 0.63 | 0.62 | 3.17E-01 | 1.07 | | | |
| rs4441471 | 2:16534140 | G/A | | 0.76 | 0.70 | 1.26E-06 | 1.43 | 4.51E-09 | 1.31 | 0.8084 |
| | | | | 0.73 | 0.70 | 1.61E-02 | 1.18 | | | |
| rs10196016 | 2:16534582 | A/G | | 0.76 | 0.69 | 1.15E-06 | 1.43 | 3.71E-09 | 1.31 | 0.8166 |
| | | | | 0.73 | 0.70 | 1.64E-02 | 1.18 | | | |
| rs10856790 | 2:16534774 | T/C | | 0.76 | 0.69 | 1.22E-06 | 1.43 | 4.69E-09 | 1.31 | 0.8224 |

| | | | | | | | | | | |
|---|---|---|---|---|---|---|---|---|---|---|
| | | | | 0.74 | 0.70 | 1.83E-02 | 1.18 | | | |
| rs10198582 | 2:16534820 | A/G | | 0.76 | 0.69 | 5.53E-06 | 1.41 | 1.28E-08 | 1.30 | 0.7669 |
| | | | | 0.73 | 0.70 | 1.51E-02 | 1.18 | | | |
| rs4832649 | 2:16535124 | C/T | | 0.76 | 0.70 | 1.58E-06 | 1.43 | 4.99E-09 | 1.31 | 0.8003 |
| | | | | 0.74 | 0.70 | 1.53E-02 | 1.18 | | | |
| rs4832650 | 2:16535131 | G/A | | 0.76 | 0.70 | 2.04E-06 | 1.43 | 6.44E-09 | 1.30 | 0.7983 |
| | | | | 0.74 | 0.70 | 1.61E-02 | 1.18 | | | |
| rs6706305 | 2:16535483 | G/A | | 0.76 | 0.70 | 1.03E-06 | 1.44 | 3.43E-09 | 1.31 | 0.8123 |
| | | | | 0.74 | 0.70 | 1.55E-02 | 1.18 | | | |
| rs6706417 | 2:16535592 | C/A | | 0.76 | 0.70 | 1.53E-06 | 1.43 | 5.18E-09 | 1.31 | 0.8062 |
| | | | | 0.74 | 0.70 | 1.63E-02 | 1.18 | | | |
| rs6713976 | 2:16536369 | G/T | | 0.76 | 0.69 | 9.13E-07 | 1.43 | 3.63E-09 | 1.31 | 0.8166 |
| | | | | 0.73 | 0.70 | 1.71E-02 | 1.18 | | | |
| rs7593098 | 2:16537037 | G/C | | 0.76 | 0.69 | 2.77E-06 | 1.42 | 8.27E-09 | 1.30 | 0.7910 |
| | | | | 0.73 | 0.70 | 1.64E-02 | 1.18 | | | |
| rs2067702 | 2:16537626 | G/A | | 0.68 | 0.62 | 1.94E-03 | 1.29 | 4.19E-05 | 1.19 | 0.8452 |
| | | | | 0.64 | 0.63 | 2.83E-01 | 1.07 | | | |
| rs4832651 | 2:16537738 | G/A | | 0.77 | 0.71 | 1.26E-06 | 1.44 | 2.60E-09 | 1.32 | 0.7857 |
| | | | | 0.75 | 0.71 | 1.12E-02 | 1.20 | | | |
| rs12623437 | 2:16538411 | G/T | | 0.77 | 0.71 | 2.27E-06 | 1.43 | 5.05E-09 | 1.31 | 0.7857 |
| | | | | 0.75 | 0.71 | 1.31E-02 | 1.19 | | | |
| rs6741506 | 2:16539467 | C/G | | 0.67 | 0.61 | 2.37E-03 | 1.29 | 7.16E-05 | 1.19 | 0.8681 |
| | | | | 0.63 | 0.61 | 3.65E-01 | 1.06 | | | |
| rs7571487 | 2:16543446 | C/T | | 0.77 | 0.70 | 1.00E-06 | 1.44 | 2.00E-09 | 1.32 | 0.8041 |
| | | | | 0.75 | 0.71 | 1.25E-02 | 1.19 | | | |
| rs7563089 | 2:16543457 | T/C | | 0.76 | 0.69 | 3.13E-06 | 1.42 | 1.12E-08 | 1.30 | 0.8063 |
| | | | | 0.73 | 0.70 | 2.09E-02 | 1.17 | | | |
| rs79434472 | 2:16543530 | T/G | | 0.67 | 0.61 | 2.62E-03 | 1.29 | 7.98E-05 | 1.18 | 0.8632 |
| | | | | 0.63 | 0.61 | 3.65E-01 | 0.94 | | | |
| rs4488663 | 2:16543538 | G/A | | 0.76 | 0.69 | 1.45E-06 | 1.43 | 5.78E-09 | 1.30 | 0.8315 |
| | | | | 0.73 | 0.70 | 2.09E-02 | 1.17 | | | |
| rs4240231 | 2:16543870 | C/T | | 0.76 | 0.69 | 3.10E-06 | 1.42 | 1.34E-08 | 1.30 | 0.8029 |
| | | | | 0.73 | 0.70 | 2.09E-02 | 1.17 | | | |
| rs4240232 | 2:16543937 | T/C | | 0.77 | 0.71 | 2.00E-06 | 1.43 | 3.49E-09 | 1.31 | 0.7693 |
| | | | | 0.75 | 0.71 | 1.03E-02 | 1.20 | | | |
| rs4832652 | 2:16544127 | A/T | | 0.76 | 0.69 | 2.86E-06 | 1.42 | 1.26E-08 | 1.30 | 0.8006 |
| | | | | 0.73 | 0.70 | 2.01E-02 | 1.18 | | | |
| rs4832653 | 2:16544281 | C/T | | 0.76 | 0.69 | 2.23E-06 | 1.42 | 1.11E-08 | 1.30 | 0.8087 |
| | | | | 0.73 | 0.70 | 2.09E-02 | 1.17 | | | |
| rs4832654 | 2:16544376 | T/C | | 0.76 | 0.69 | 2.71E-06 | 1.42 | 1.27E-08 | 1.30 | 0.8095 |
| | | | | 0.73 | 0.70 | 2.20E-02 | 1.17 | | | |
| rs4832655 | 2:16544419 | A/G | | 0.76 | 0.69 | 3.03E-06 | 1.42 | 6.66E-09 | 1.30 | 0.7729 |
| | | | | 0.74 | 0.70 | 1.27E-02 | 1.19 | | | |

| | | | | | | | | | |
|---|---|---|---|---|---|---|---|---|---|
| rs59197172[f] | 2:16545035 | -/AT | 0.76 | 0.69 | 2.54E-06 | 1.42 | - | - | - |
| | | | 0.73 | 0.70 | 2.01E-02 | 1.18 | | | |
| rs4832657 | 2:16545471 | T/C | 0.68 | 0.62 | 1.56E-03 | 1.30 | 3.75E-05 | 1.19 | 0.8572 |
| | | | 0.64 | 0.63 | 2.99E-01 | 1.07 | | | |
| rs4832658 | 2:16545663 | T/C | 0.67 | 0.61 | 1.62E-03 | 1.29 | 5.44E-05 | 1.19 | 0.8729 |
| | | | 0.63 | 0.61 | 3.65E-01 | 1.06 | | | |
| rs5829552[f] | 2:16545695 | A/- | 0.76 | 0.69 | 9.48E-07 | 1.44 | - | - | - |
| | | | 0.73 | 0.70 | 2.44E-02 | 1.17 | | | |
| rs12613096 | 2:16546002 | C/G | 0.77 | 0.70 | 1.16E-06 | 1.44 | 2.46E-09 | 1.32 | 0.7912 |
| | | | 0.75 | 0.71 | 1.14E-02 | 1.20 | | | |
| rs4441472 | 2:16546584 | A/G | 0.76 | 0.69 | 2.65E-06 | 1.42 | 1.22E-08 | 1.30 | 0.8046 |
| | | | 0.73 | 0.70 | 2.08E-02 | 1.17 | | | |
| rs4319933 | 2:16546763 | G/A | 0.76 | 0.69 | 3.27E-06 | 1.42 | 1.52E-08 | 1.29 | 0.8045 |
| | | | 0.73 | 0.70 | 2.19E-02 | 1.17 | | | |
| rs5829553[f] | 2:16546890 | TAACT/- | 0.77 | 0.71 | 1.50E-06 | 1.44 | - | - | - |
| | | | 0.75 | 0.71 | 1.52E-02 | 1.19 | | | |
| rs4303719 | 2:16547251 | T/G | 0.76 | 0.69 | 4.32E-06 | 1.41 | 1.67E-08 | 1.29 | 0.7949 |
| | | | 0.73 | 0.70 | 2.02E-02 | 1.18 | | | |
| rs4240233 | 2:16547342 | A/G | 0.77 | 0.71 | 1.90E-06 | 1.43 | 3.53E-09 | 1.31 | 0.7827 |
| | | | 0.75 | 0.71 | 1.15E-02 | 1.19 | | | |
| rs7566780 | 2:16548089 | G/A | 0.76 | 0.69 | 2.72E-06 | 1.42 | 1.14E-08 | 1.30 | 0.8035 |
| | | | 0.73 | 0.70 | 1.99E-02 | 1.18 | | | |
| rs7566781 | 2:16548090 | C/A | 0.76 | 0.69 | 2.34E-06 | 1.42 | 1.00E-08 | 1.30 | 0.8074 |
| | | | 0.73 | 0.70 | 1.99E-02 | 1.18 | | | |
| rs10168963 | 2:16548142 | C/A | 0.67 | 0.61 | 2.34E-06 | 1.42 | 1.08E-04 | 1.18 | 0.8576 |
| | | | 0.63 | 0.61 | 3.83E-01 | 1.06 | | | |
| rs7570487 | 2:16549078 | T/A | 0.77 | 0.70 | 2.94E-06 | 1.42 | 2.07E-08 | 1.30 | 0.8285 |
| | | | 0.74 | 0.71 | 3.15E-02 | 1.16 | | | |
| rs4240234 | 2:16551007 | T/G | 0.76 | 0.69 | 2.16E-06 | 1.42 | 7.44E-09 | 1.30 | 0.7982 |
| | | | 0.73 | 0.70 | 1.72E-02 | 1.18 | | | |
| rs4263114 | 2:16551123 | G/T | 0.76 | 0.70 | 4.29E-06 | 1.42 | 1.12E-08 | 1.30 | 0.7817 |
| | | | 0.73 | 0.70 | 1.63E-02 | 1.18 | | | |
| rs10172734 | 2:16551786 | A/G | 0.76 | 0.69 | 3.06E-06 | 1.42 | 2.38E-08 | 1.29 | 0.8295 |
| | | | 0.73 | 0.70 | 3.26E-02 | 1.16 | | | |
| rs5007483 | 2:16552257 | T/G | 0.68 | 0.62 | 4.71E-03 | 1.28 | 1.83E-04 | 1.17 | 0.8683 |
| | | | 0.63 | 0.62 | 4.58E-01 | 1.05 | | | |
| rs76649769[f] | 2:16552304 | -/AGG | 0.66 | 0.61 | 2.75E-03 | 1.28 | - | - | - |
| | | | 0.62 | 0.61 | 4.99E-01 | 1.05 | | | |
| rs7552 | 2:16552660 | G/A | 0.77 | 0.71 | 1.76E-06 | 1.43 | 2.18E-09 | 1.32 | 0.7667 |
| | | | 0.75 | 0.71 | 9.01E-03 | 1.20 | | | |
| rs6743149 | 2:16553779 | C/T | 0.77 | 0.70 | 3.95E-06 | 1.42 | 2.99E-09 | 1.31 | 0.7135 |
| | | | 0.74 | 0.71 | 6.20E-03 | 1.21 | | | |
| rs5829555[f] | 2:16555361 | A/- | 0.76 | 0.70 | 1.67E-05 | 1.39 | - | - | - |

| | | | | |
|---|---|---|---|---|
| | 0.74 | 0.71 | 7.98E-03 | 1.21 |

[a] Discovery: targeted sequencing in 1,414 NSCLP cases and 1,371 controls of Chinese; Replication: replication study in 1,025 NSCLP cases and 1,022 controls of Chinese.

[b] F_A, allele frequency in cases; [c] F_U, allele frequency in controls; [d] OR: odds ratio.

[e] $P_{\mathrm{HET}}$: P-value for Heterogeneity, $P_{\mathrm{HET}} > 0.05$ was considered to signify no heterogeneity.

[f] Short insertions and deletions (Indels) were excluded from the pooled meta-analysis due to allele format incompatibilities with the meta-analysis algorithm (METAL).

**Table S3: Potential enhancer fragments detected by genetic screening.**

| Name | Location(hg38) | Included SNPs |
|---|---|---|
| Enh1 | 2:16509542-16510157 | rs34317246 |
| Enh2 | 2:16510223-16511157 | rs11096594 |
| Enh3 | 2:16512322-16512831 | rs4564771 |
| Enh4 | 2:16513938-16514332 | rs35165397 |
| Enh5 | 2:16515185-16515750 | rs7560047 |
| Enh6 | 2:16516635-16516950 | rs6715080 |
| Enh7 | 2:16517940-16518310 | rs7570255 |
| Enh8 | 2:16518750-16519371 | rs11695429 |
| Enh9 | 2:16521089-16521686 | rs4632345 |
| | | rs67530244 |
| Enh10 | 2:16523745-16524498 | rs4832647 |
| Enh11 | 2:16524318-16525120 | rs6735186 |
| Enh12 | 2:16525097-16525838 | rs4493263 |
| | | rs4240230 |
| | | rs4497881 |
| Enh13 | 2:16525946-16526917 | rs12710725 |
| | | rs13406059 |
| Enh14 | 2:16526902-16527551 | rs4832465 |
| Enh15 | 2:16527373-16528309 | rs4832648 |
| Enh16 | 2:16529201-16529410 | rs4832468 |
| Enh17 | 2:16530413-16531416 | rs11897201 |
| Enh18 | 2:16533769-16534575 | rs4441471 |
| Enh19 | 2:16534213-16534855 | rs10196016 |
| | | rs10856790 |
| | | rs10198582 |
| Enh20 | 2:16534833-16535393 | rs4832649 |
| | | rs4832650 |
| Enh21 | 2:16535147-16535710 | rs6706305 |
| | | rs6706417 |
| Enh22 | 2:16536043-16536668 | rs6713976 |
| Enh23 | 2:16536593-16537162 | rs7593098 |
| Enh24 | 2:16537465-16537939 | rs4832651 |
| Enh25 | 2:16537933-16538721 | rs12623437 |
| Enh26 | 2:16543262-16543860 | rs7571487 |
| | | rs7563089 |
| | | rs4488663 |
| Enh27 | 2:16543553-16544110 | rs4240231 |
| | | rs4240232 |
| Enh28 | 2:16543938-16544640 | rs4832652 |
| | | rs4832653 |

| | | |
|---|---|---|
| | | rs4832654 |
| | | rs4832655 |
| Enh29 | 2:16545874-16546387 | rs12613096 |
| Enh30 | 2:16546434-16547054 | rs4441472 |
| | | rs4319933 |
| Enh31 | 2:16547101-16547452 | rs4303719 |
| | | rs4240233 |
| Enh32 | 2:16547914-16548523 | rs7566780 |
| | | rs7566781 |
| Enh33 | 2:16548874-16549245 | rs7570487 |
| Enh34 | 2:16550729-16551369 | rs4240234 |
| | | rs4263114 |
| Enh35 | 2:16551519-16552252 | rs10172734 |
| Enh36 | 2:16552534-16553040 | rs7552 |
| Enh37 | 2:16553560-16554034 | rs6743149 |
| C1 | 2:16516950-16517749 | - |

**Table S4: The Haplotype of candidate function SNPs residing within Enh34 in NSCLP cohort.**

| SNPs | Haplotype | case_F | control_F | total_group_F | Pvalue |
|---|---|---|---|---|---|
| rs4240234, rs4263114 | TG | 2145(76.06%) | 1892(69.30%) | 4037(72.74%) | 5.80E-08 |
| rs4240234, rs4263114 | GT | 671(23.79%) | 829(30.37%) | 1500(27.03%) | |

**Table S5: Zebrafish-based *in vivo* assay for C1 and Enh-*MYCN* with different alleles.**

| Element | Replicate | Total injected | pattern | | |
|---|---|---|---|---|---|
| | | | strong | weak | none |
| Control_C1 | replicate_1 | 184 | 13 | 28 | 143 |
| | replicate_2 | 203 | 22 | 31 | 150 |
| | replicate_3 | 191 | 19 | 35 | 137 |
| Enh-*MYCN* (rs4263114_T) | replicate_1 | 196 | 97 | 53 | 46 |
| | replicate_2 | 187 | 88 | 47 | 52 |
| | replicate_3 | 210 | 103 | 51 | 56 |
| rs4263114_G | replicate_1 | 188 | 31 | 49 | 108 |
| | replicate_2 | 175 | 27 | 44 | 104 |
| | replicate_3 | 193 | 39 | 55 | 99 |

**Table S6: The candidate transcription factors predicted to exhibit dynamic binding activity around linked SNPs.**

| TF | SNP bound | Risk allele/non-risk allele | Allele preferred | Predicted capacity in *MYCN* expression | Matrix ID | Database |
|---|---|---|---|---|---|---|
| FOXP1 | rs4263114 | G/T | T | upregulation | MA0481.3 | JASPAR |
| FOXP2 | rs4263114 | G/T | T | upregulation | MA0593.1 | JASPAR |
| HOXB2 | rs4263114 | G/T | T | upregulation | MA0902.1 | JASPAR |
| SOX4 | rs4263114 | G/T | G | downregulation | MA0867.3 | JASPAR |
| VAX2 | rs4263114 | G/T | T | upregulation | MA0723.3 | JASPAR |
| VDR | rs4263114 | G/T | G | downregulation | MA0693.2 | JASPAR |
| AR | rs4240234 | T/G | T | downregulation | MA0007.2 | JASPAR/hTFtarget |
| MSC | rs4240234 | T/G | G | upregulation | MA0665.1 | JASPAR |
| PPARG | rs4240234 | T/G | G | upregulation | MA0066.1 | AnimalTFDB |
| TCF7L2 | rs4240234 | T/G | G | upregulation | MA0523.1 | JASPAR |
| ZBTB12 | rs4240234 | T/G | T | downregulation | MA1649.1 | JASPAR |
| ZBTB26 | rs4240234 | T/G | T | downregulation | MA1579.2 | JASPAR |

**Table S7: Oligos used in the study.**

| Application | Name | Sequence |
|---|---|---|
| plasmids construction | pGL6-*MYCN*pro-F | aaacttggtctgacagcggccgcTCAAGCTGTTTGAGCCGAGG |
| plasmids construction | pGL6-*MYCN*pro-R | aaacttggtctgacagcggccgcTCAAGCTGTTTGAGCCGAGG |
| plasmids construction | E1-F | ctggcctaactggccggtaccAACCTGCTCCCCAAAGCTACC |
| plasmids construction | E1-R | gctcaaacagcttgagctagcCTGGCAGAATACCTGGCACAT |
| plasmids construction | E2-F | ctggcctaactggccggtaccATGCATCAGACTGGTAGGGACTATT |
| plasmids construction | E2-R | gctcaaacagcttgagctagcTCTAGATGGCCCAGGGATCTT |
| plasmids construction | E3-F | ctggcctaactggccggtaccGCTTAGACCTCTGGGCACTGG |
| plasmids construction | E3-R | gctcaaacagcttgagctagcCTTGTGCCCAGCCTGAAAAC |
| plasmids construction | E4-F | ctggcctaactggccggtaccGCAAACAAGTGATTGCAAACCA |
| plasmids construction | E4-R | gctcaaacagcttgagctagcAGGGCAGACCTTGCCTAGTATATT |
| plasmids construction | E5-F | ctggcctaactggccggtaccGGTGCCAGATAAATAGAAGGGGT |
| plasmids construction | E5-R | gctcaaacagcttgagctagcGGCTTCCCTGGGCTTACATT |
| plasmids construction | E6-F | ctggcctaactggccggtaccAGGTGCTATTGTTCTAGAAGGAGTTTT |
| plasmids construction | E6-R | gctcaaacagcttgagctagcCAGTCTGTGAGTCACGTTTGAACC |
| plasmids construction | E7-F | gagcttcaaagcgcaagatctCACCTCACAGGCTAGTCTCAAATAA |
| plasmids construction | E7-R | taccctctagtgtctaagcttTAAGGCTGCAGCTCACTCAACA |
| plasmids construction | E8-F | ctggcctaactggccggtaccGCCAGTCATAGGCAGCTAGGG |
| plasmids construction | E8-R | gctcaaacagcttgagctagcCATCCAGGGTCCTTGTGTCCT |
| plasmids construction | E9-F | ctggcctaactggccggtaccAATGGGGTGATGGTGCCAA |
| plasmids construction | E9-R | gctcaaacagcttgagctagcTGCAAGGAGTCAAGAGCCTGG |

| | | |
|---|---|---|
| plasmids construction | E10-F | ctggcctaactggccggtaccTCTCCAGGACCGTGAATTGC |
| plasmids construction | E10-R | gctcaaacagcttgagctagcTTGCCGGTCTAGACGAAGAGA |
| plasmids construction | E11-F | ctggcctaactggccggtaccCCCCATCTTTGCCCAGCT |
| plasmids construction | E11-R | gctcaaacagcttgagctagcACTGAACTCGCTGCCTTTCATT |
| plasmids construction | E12-F | gagcttcaaagcgcaagatctCCAATGAAAGGCAGCGAGTT |
| plasmids construction | E12-R | taccctctagtgtctaagcttCGCCTCAGATACAGGTAGGCTG |
| plasmids construction | E13-F | ctggcctaactggccggtaccCATCCAGCAGAGTGCAAACCC |
| plasmids construction | E13-R | gctcaaacagcttgagctagcCCCAGCAACACTGGGTAAGTTT |
| plasmids construction | E14-F | ctggcctaactggccggtaccACCCAGTGTTGCTGGGTAATCA |
| plasmids construction | E14-R | gctcaaacagcttgagctagcTGGGAAATGGTGTGCAGGTT |
| plasmids construction | E15-F | ctggcctaactggccggtaccAGTAATTGTGACAGAGATCACGTGG |
| plasmids construction | E15-R | gctcaaacagcttgagctagcCTAAGAGTTAGAGTTAAGCTAAGACCCTACC |
| plasmids construction | E16-F | ctggcctaactggccggtaccGAGTGTGCCATATTGACCTATTGTCA |
| plasmids construction | E16-R | gctcaaacagcttgagctagcCTTCCACAGATGCTTTCCAGATG |
| plasmids construction | E17-F | ctggcctaactggccggtaccAGTTGGCAGGAGATGTGTGGG |
| plasmids construction | E17-R | gctcaaacagcttgagctagcGAAAGGGGCAGACATTCCCA |
| plasmids construction | E18-F | ctggcctaactggccggtaccTGCAGGAAACTGAACCAACCTT |
| plasmids construction | E18-R | gctcaaacagcttgagctagcAAGTAGTGTGGTGGTAGCCCTGTA |
| plasmids construction | E19-F | ctggcctaactggccggtaccCAATCCCCCATGCCTCATC |
| plasmids construction | E19-R | gctcaaacagcttgagctagcGCTACAGTGCAGGCAGACTCTTT |
| plasmids construction | E20-F | ctggcctaactggccggtaccAAGGCCTGGCGAATATGCA |
| plasmids | E20-R | gctcaaacagcttgagctagcCCACCACGGAGGCTAG |

| | | |
|---|---|---|
| construction | | AAACT |
| plasmids construction | E21-F | ctggcctaactggccggtaccGCTGCACCATTCACAG GTTGA |
| plasmids construction | E21-R | gctcaaacagcttgagctagcGCCCACCATGTGTAGA CAGTATATG |
| plasmids construction | E22-F | ctggcctaactggccggtaccACAGCACAATGATTGA GAGCACA |
| plasmids construction | E22-R | gctcaaacagcttgagctagcCCTCTATCAGAAACAG GCGGC |
| plasmids construction | E23-F | ctggcctaactggccggtaccTCCTCTCTAAAAATGGA AGACTGGA |
| plasmids construction | E23-R | gctcaaacagcttgagctagcTGATTGTTCTGGCTGCT AGGTTG |
| plasmids construction | E24-F | ctggcctaactggccggtaccGATTTCTTGACTTTCCC CCAACC |
| plasmids construction | E24-R | gctcaaacagcttgagctagcGCCGTGAATGGCATGTT GA |
| plasmids construction | E25-F | ctggcctaactggccggtaccTCACGGCTGCTTCTCA AACA |
| plasmids construction | E25-R | gctcaaacagcttgagctagcGCACTGAGGGGAAGCT CAATAC |
| plasmids construction | E26-F | ctggcctaactggccggtaccTGGTTGTTTGAGAATG GCAGG |
| plasmids construction | E26-R | gctcaaacagcttgagctagcGGATTGATCCCATCCAC TGGC |
| plasmids construction | E27-F | ctggcctaactggccggtaccTACACCTGCCCCACTCT CATATG |
| plasmids construction | E27-R | gctcaaacagcttgagctagcTTGGGCTGGTTGGCCTA AG |
| plasmids construction | E28-F | ctggcctaactggccggtaccGGTTGTTGACCCAGGA ACTTTG |
| plasmids construction | E28-R | gctcaaacagcttgagctagcTCTGGCATGGACTGTG CTATCC |
| plasmids construction | E29-F | ctggcctaactggccggtaccTGAGTGTTGGCTGTGT GGAGAA |
| plasmids construction | E29-R | gctcaaacagcttgagctagcGGCACCAGAAGCAGAG AAGTTC |
| plasmids construction | E30-F | ctggcctaactggccggtaccGCCTTGTCCCAGACTG ATGTCC |
| plasmids construction | E30-R | gctcaaacagcttgagctagcCCACACCTATGGTGCTG TCATTA |
| plasmids construction | E31-F | ctggcctaactggccggtaccAGAGTTCACCAGAGCT CCCTAACA |

| | | |
|---|---|---|
| plasmids construction | E31-R | gctcaaacagcttgagctagcTGTTTGCACAGTGACATGCTTTG |
| plasmids construction | E32-F | ctggcctaactggccggtaccTCAATTTTCAAAAAGAGTGTCTCCA |
| plasmids construction | E32-R | gctcaaacagcttgagctagcGGAGCTCAGCTGACCACATTCA |
| plasmids construction | E33-F | ctggcctaactggccggtaccTGGAAGGTCTCTCTTGGGACTTC |
| plasmids construction | E33-R | gctcaaacagcttgagctagcCCCTGAATAACAGCCAAAGGC |
| plasmids construction | E34-F | ctggcctaactggccggtaccGACATGGGTTCAAGCCCCTC |
| plasmids construction | E34-R | gctcaaacagcttgagctagcAGAAGGAGTCACCCAGGCATT |
| plasmids construction | E35-F | ctggcctaactggccggtaccGGAGCCATCCTGTATGACCACA |
| plasmids construction | E35-R | acagcttgagctagcggtaccGCCACTGGACTGATGAAGGTG |
| plasmids construction | E36-F | ctggcctaactggccggtaccTCTTGTAGCTGAGCCGACACAG |
| plasmids construction | E36-R | gctcaaacagcttgagctagcCACTGTTTGCCGCTAACCTCTA |
| plasmids construction | E37-F | ctggcctaactggccggtaccATGGGCGCTCCATGTTAAGTT |
| plasmids construction | E37-R | gctcaaacagcttgagctagcACCCTGAGGTTGATGCTGTTTT |
| plasmids construction | C1-F | ctggcctaactggccggtaccGGTCTCTTTAACTTGCCAAGCAA |
| plasmids construction | C1-R | cgtgagctcctcgaggctagcAATGCTGCTCTATACTAGAAACCACAC |
| plasmids construction | rs35165397mutantE4-F | GATTAGTGTAAATTtTTTTATTCTGATTTAATCTCAAATTAAAGC |
| plasmids construction | rs35165397mutantE4-R | AAaAATTTACACTAATCTTTCCTGCAAAAAAG |
| plasmids construction | rs4632345mutantE9-F | CAgTCATTAAGTCCTCCAGAGCTTTCTT |
| plasmids construction | rs4632345mutantE9-R | GGAGGACTTAATGAcTGATTATATGTGGGGAGAAAAGTCA |
| plasmids construction | rs67530244mutantE9-F | ATCTGAGGCTGGAgGAAGGTTCACGGCACTTTCCT |
| plasmids construction | rs67530244mutantE9-R | TTCcTCCAGCCTCAGATGAAGAAAAGA |
| plasmids | double mutantE9-F | ccagagctttcttctgcattctgtggggctgcccccaGAAGGTT |

| | | |
|---|---|---|
| construction | | CACGGCACTTTCCT |
| plasmids construction | double mutantE9-R | tgcagaagaaagctctggaggacttaatgacTGATTATATGTGGGGAGAAAAGTCA |
| plasmids construction | rs4240234mutantE34-F | AGtTCTTGAAGACCACCTAGCATATTTCT |
| plasmids construction | rs4240234mutantE34-R | GGTGGTCTTCAAGAaCTTTTTGAACCTACAGATTCTAAACCTT |
| plasmids construction | rs4623114mutantE34-F | CCTTCTGTTGACTAAgGTTTAGATGCAGACCAGGGAATCT |
| plasmids construction | rs4623114mutantE34-R | CcTTAGTCAACAGAAGGAAAAGGCTCT |
| plasmids construction | double mutantE34-F | cttgaagaccacctagcatatttctattctgatgagagaaactccatcGTTTAGATGCAGACCAGGGAATCT |
| plasmids construction | double mutantE34-R | tgctaggtggtcttcaagaaCTTTTTGAACCTACAGATTCTAAACCTT |
| 3C PCR | P-P1 | TGATGATGACCACGACATGC |
| 3C PCR | P-P2 | AAGGCATCCTGGCTAGAGGA |
| 3C PCR | P-P3 | CCATGTTCTGACCAGTGGTA |
| 3C PCR | P-P4 | TAGGAAGCTGTAATCAGCCT |
| 3C PCR | P-E1 | AAGCCAAGACTCAGCAGTAC |
| 3C PCR | P-E2 | GAGTGCACCTCTGAGGACCT |
| genome editing | Enh-*MYCN*-KO-gRNA1 | GCCACCACAGGGCTCATGAGGGG |
| genome editing | Enh-*MYCN*-KO-gRNA2 | GCCTGGGTGACTCCTTCTAGTGG |
| genome editing | KO validation-F1 | CTGTTAGTGATCACAGACATTCAG |
| genome editing | KO validation-F2 | CCTGGTATGATCACATATTCTGG |
| genome editing | KO validation-F3 | GTGACTGCCAAATAGGATATC |
| genome editing | KO validation-R1 | CTTAGTACGTAGTATCCAGCAAG |
| genome editing | rs4263114mutant-gRNA | TCTAAACATTAGTCAACAGAAGG |
| genome editing | Oligo1 Sequence | ATCCAAGCAGCTGTACTTTTTCAACTTGAAAATCTCCAAGGAAAAGAGCCTTTTCCTTCTGTTGACTAAGGTTTAGATGCAGACCAGGGAATCTTTCCTGTCACTTGGCTGAACTCCCTTGCTAAAACAA |
| genome editing | mutant validation-F1 | TTCCACTAGAAGGAGTCACCCA |
| genome editing | mutant validation-R1 | AGAGACACACGACATGGGTTC |
| RT-qPCR | MYCN-Homo-qp-F | GACTGTAGCCATCCGAGGAC |
| RT-qPCR | MYCN-Homo-qp-R | ATCTTCATCATCTCCCGCCG |
| RT-qPCR | Homo-GAPDH-qPCR-F | GCACCGTCAAGGCTGAGAAC |
| RT-qPCR | Homo-GAPDH-qPCR-R | TGGTGAAGACGCCAGTGGA |
| RT-qPCR | Homo-qp-SP7-F | CCTCTGCGGGACTCAACAAC |
| RT-qPCR | Homo-qp-SP7-R | AGCCCATTAGTGCTTGTAAAGG |
| RT-qPCR | Homo-qp-ALP(ALPL)-F | GCTGTAAGGACATCGCCTACCA |
| RT-qPCR | Homo-qp-ALP(ALPL)-R | CCTGGCTTTCTCGTCACTCTCA |

| | | |
|---|---|---|
| RT-qPCR | Homo-qp-RUNX2-F | GCGCATTCCTCATCCCAGTA |
| RT-qPCR | Homo-qp-RUNX2-R | GGCTCAGGTAGGAGGGGTAA |
| RT-qPCR | Homo-qp-COL1a1-F | TTTGGATGGTGCCAAGGGAG |
| RT-qPCR | Homo-qp-COL1a1-R | AGTAGCACCATCATTTCCACGA |
| RT-qPCR | Homo-qp-OCN-F | AGCGAGGTAGTGAAGAGAC |
| RT-qPCR | Homo-qp-OCN-R | GAAAGCCGATGTGGTCAG |
| RT-qPCR | Homo-qp-SOX2-F | GCTACAGCATGATGCAGGACCA |
| RT-qPCR | Homo-qp-SOX2-R | TCTGCGAGCTGGTCATGGAGTT |
| RT-qPCR | Homo-qp-OCT4-F | GACAGGGGGAGGGGAGGAGCTAGG |
| RT-qPCR | Homo-qp-OCT4-R | CTTCCCTCCAACCAGTTGCCCCAAAC |
| RT-qPCR | Homo-qp-NANOG-F | CAGCCCAGATTCTTCCACCAGTCCC |
| RT-qPCR | Homo-qp-NANOG-R | CGGAAGCTTCCCAGTCGGGTTCAC |
| RT-qPCR | Homo-qp-p75-F | CTGCCTGGACAGCGTGACGTT |
| RT-qPCR | Homo-qp-p75-R | GCAGCGCCCAGTCGTCTCAT |
| RT-qPCR | Homo-qp-SOX10-F | CTCAGCGGCTACGACTGGA |
| RT-qPCR | Homo-qp-SOX10-R | GGCGCTTGTCACTTTCGTTC |
| RT-qPCR | Homo-qp-AP2α-F | AGGTCAATCTCCCTACACGAG |
| RT-qPCR | Homo-qp-AP2α-R | GGAGTAAGGATCTTGCGACTGG |
| RT-qPCR | Homo-qp-HOXA1-F | TCCTGGAATACCCCATACTTAGC |
| RT-qPCR | Homo-qp-HOXA1-R | GCACGACTGGAAAGTTGTAATCC |
| RT-qPCR | Homo-qp-HOXA2-F | GCATCACCAAACAAAAACTCCTTTG |
| RT-qPCR | Homo-qp-HOXA2-R | TGGGTCAGGGAATCACTAAACAGAA |
| RT-qPCR | Homo-qp-HOXA3-F | ACCAACTAACGCCTAAACCTCGGTA |
| RT-qPCR | Homo-qp-HOXA3-R | GCAGATTTTTGGAGCAATTCTTTCC |
| RT-qPCR | Homo-qp-HOXB2-F | TTCACCAGTACGCTCTGTGC |
| RT-qPCR | Homo-qp-HOXB2-R | TTTTCCAGTAGACGGCCAAG |
| RT-qPCR | Homo-qp-PAX3-F | CGGCATCCTGAGCGAGCGAG |
| RT-qPCR | Homo-qp-PAX3-R | ACTCGGGCCTCGGTGAGCTT |
| RT-qPCR | Homo-qp-DDX1-F | AAACCAAGCCCTCTTTCCTGCC |
| RT-qPCR | Homo-qp-DDX1-R | GCCTTGGAAAGAGCAACAAAGCC |
| RT-qPCR | FOXP1-Homo-qp-F | CAAAGAACGCCTGCAAGCCATG |
| RT-qPCR | FOXP1-Homo-qp-R | GGAGTATGAGGTAAGCTCTGTGG |
| RT-qPCR | FOXP2-Homo-qp-F | TGGATGACCGAAGCACTGCTCA |
| RT-qPCR | FOXP2-Homo-qp-R | TGGGAGATGGTTTGGGCTCTGA |
| RT-qPCR | TCF7L2-Homo-qp-F | GAATCGTCCCAGAGTGATGTCG |
| RT-qPCR | TCF7L2-Homo-qp-R | TGCACTCAGCTACGACCTTTGC |
| RT-qPCR | MSC-Homo-qp-F | AGGACCGCTATGAGAACGGCTA |
| RT-qPCR | MSC-Homo-qp-R | GTGGTTCCACATAGTCTGTTGGC |
| RT-qPCR | PDPN-Homo-qp-F | GTGCCGAAGATGATGTGGTGAC |
| RT-qPCR | PDPN-Homo-qp-R | GGACTGTGCTTTCTGAAGTTGGC |
| RT-qPCR | VAX2-Homo-qp-F | GGACACGTACATCCTTCACTGC |
| RT-qPCR | VAX2-Homo-qp-R | GGTTCTGGAACCAGACCTTCAC |
| RT-qPCR | CEBPD-Homo-qp-F | TCCGGCAGTTCTTCAAGCAGCT |

| | | |
|---|---|---|
| RT-qPCR | CEBPD-Homo-qp-R | GAGGTATGGGTCGTTGCTGAGT |
| RT-qPCR | ZBTB26-Homo-qp-F | GAGCCACATTGTAGAACGGTGC |
| RT-qPCR | ZBTB26-Homo-qp-R | GGAGAAGCACTCTGTGGTTCAC |
| RT-qPCR | ZBTB12-Homo-qp-F | ATCTGCGGCAAGTGCTTCACAC |
| RT-qPCR | ZBTB12-Homo-qp-R | TAATGGCAGGCTTGTGGGCGAA |
| RT-qPCR | SOX4-Homo-qp-F | CTCTCCAGCCTGGGAACTATAA |
| RT-qPCR | SOX4-Homo-qp-R | CGGAGGTGGGTAAAGAGAGAA |
| RT-qPCR | AR-Homo-qp-F | ATGGTGAGCAGAGTGCCCTATC |
| RT-qPCR | AR-Homo-qp-R | ATGGTCCCTGGCAGTCTCCAAA |
| RT-qPCR | VDR-Homo-qP-F | CGCATCATTGCCATACTGCTGG |
| RT-qPCR | VDR-Homo-qP-R | CCACCATCATTCACACGAACTGG |
| RT-qPCR | PPARG-Homo-qP-F | AGCCTGCGAAAGCCTTTTGGTG |
| RT-qPCR | PPARG-Homo-qP-R | GGCTTCACATTCAGCAAACCTGG |
| ChIP-qPCR | DNA fragment covering rs4263114-F | CCATCCAAGCAGCTGTACTTTTTC |
| ChIP-qPCR | DNA fragment covering rs4263114-R | GCTGATCTTCCCTTGGATCCTC |
| ChIP-qPCR | off-target control-F | TTGTTTTCCATTGTTAGGTTAGTCA |
| ChIP-qPCR | off-target control-R | TGGTCCACCTAATGCAAAAACG |
| ChIP-qPCR | FOXP2 positive control-F | AAGAAACCAGGAATCCGCCC |
| ChIP-qPCR | FOXP2 positive control-R | AAATATAGGCCGGCGCGAAC |
| ChIP-qPCR | FOXP1 positive control-F | GGCTGAGCGGTGGATGTC |
| ChIP-qPCR | FOXP1 positive control-R | CACCAGCTCTCACGCCAAAA |

168 **Table S8: Antibodies used in the study.**

| Application | Antigen | Host | Dilution | Company | Cat. No. |
|---|---|---|---|---|---|
| FACS | anti-human CD271(p75) PE-CY7 | mouse | 5ul/test | BD | 562122 |
| FACS | anti-human CD57(HNK1) BV605 | mouse | 5ul/test | BD | 567205 |
| Immunofluorescence | ALP | rabbit | 1:100 | Abclonal | A0514 |
| Immunofluorescence | Ap2α | mouse | 1:200 | 3B5 | DSHB |
| Immunofluorescence | COL1 | rabbit | 1:200 | Abcam | ab21286 |
| Immunofluorescence | DLX2 | rabbit | 1:200 | Proteintech | 26244-1-AP |
| Immunofluorescence | DLX5 | mouse | 1:200 | Proteintech | 67111 |
| Immunofluorescence | FOXP2 | Goat | 1:200 | Novus | NB100-55411 |
| Immunofluorescence | HAND2 | Goat | 1:200 | R&D Systems | AF3876 |
| Immunofluorescence | OCN | rabbit | 1:200 | Proteintech | 23418-1-AP |
| Immunofluorescence | p75 | rabbit | 1:200 | Abcam | ab52987 |
| Immunofluorescence | PDPN | rabbit | 1:100 | Abclonal | A28229 |
| Immunofluorescence | SOST | rabbit | 1:100 | Abclonal | A8213 |
| Immunofluorescence | SOX10 | rabbit | 1:200 | Abcam | ab155279 |
| Immunofluorescence | SP7 | rabbit | 1:200 | Abcam | ab209484 |
| Immunofluorescence | Alexa Fluor 488 Phalloidin | | 1:200 | Thermo Fisher Scientific | A12379 |
| Immunofluorescence | Donkey anti-Goat IgG Alexa 488 | | 1:500 | Abcam | ab150129 |
| Immunofluorescence | Goat anti-mouse IgG Alexa 594 | | 1:500 | Abcam | ab150116 |
| Immunofluorescence | Goat anti-rabbit IgG Alexa 594 | | 1:500 | Abcam | ab150080 |
| Immunofluorescence | Goat anti-mouse IgG Alexa 488 | | 1:500 | Invitrogen | A28175 |
| Immunofluorescence | Goat anti-rabbit IgG Alexa 488 | | 1:500 | Invitrogen | A11008 |
| Western blotting | ALP | rabbit | 1:500 | Abclonal | A0514 |
| Western blotting | FOXP2 | Goat | 1:1000 | Novus | NB100-55411 |
| Western blotting | MYCN | mouse | 1:1000 | Abcam | ab16898 |
| Western blotting | MYCN | rabbit | 1:500 | Abclonal | A0499 |
| Western blotting | RUNX2 | rabbit | 1:500 | Abclonal | A11753 |
| Western blotting | SP7 | rabbit | 1:1000 | Abcam | ab209484 |
| Western blotting | 3×Flag | rabbit | 1:1000 | Abcam | ab205606 |
| Western blotting | GAPDH mAb-HRP-DirecT | | 1:1000 | MBL | M171-7 |
| Western blotting | HRP-labeled Donkey Anti-Goat IgG(H+L) | | 1:1000 | Beyotine | A0181 |
| Western blotting | HRP-labeled Goat Anti-Rabbit IgG(H+L) | | 1:1000 | Beyotine | A0208 |

| | | | | | |
|---|---|---|---|---|---|
| Western blotting | HRP-labeled Goat Anti-Mouse IgG(H+L) | | 1:1000 | Beyotine | A0216 |
| ChIP | FOXP1 | rabbit | 0.8μg/test | CST | 4402S |
| ChIP | FOXP2 | Goat | 0.8μg/test | Novus | NB100-55411 |
| ChIP | Normal Rabbit IgG | rabbit | 0.8μg/test | CST | 2729S |
